# omicSynth: an Open Multi-omic Community Resource for Identifying Druggable Targets across Neurodegenerative Diseases

**DOI:** 10.1101/2023.04.06.23288266

**Authors:** Chelsea X. Alvarado, Mary B. Makarious, Cory A. Weller, Dan Vitale, Mathew J. Koretsky, Sara Bandres-Ciga, Hirotaka Iwaki, Kristin Levine, Andrew Singleton, Faraz Faghri, Mike A. Nalls, Hampton L. Leonard

**Author notes:** **Correspondence** Hampton Leonard, Center for Alzheimer’s and Related Dementias, National Institute on Aging, Intramural Research Program at the National Institutes of Health Building T44, Bethesda, MD 20892, USA.

## Abstract

Treatments for neurodegenerative disorders remain rare, although recent FDA approvals, such as Lecanemab and Aducanumab for Alzheimer’s Disease, highlight the importance of the underlying biological mechanisms in driving discovery and creating disease modifying therapies. The global population is aging, driving an urgent need for therapeutics that stop disease progression and eliminate symptoms. In this study, we create an open framework and resource for evidence-based identification of therapeutic targets for neurodegenerative disease. We use Summary-data-based Mendelian Randomization to identify genetic targets for drug discovery and repurposing. In parallel, we provide mechanistic insights into disease processes and potential network-level consequences of gene-based therapeutics. We identify 116 Alzheimer’s disease, 3 amyotrophic lateral sclerosis, 5 Lewy body dementia, 46 Parkinson’s disease, and 9 Progressive supranuclear palsy target genes passing multiple test corrections (p_SMR_multi_ < 2.95×10^−6^ and p_HEIDI_ > 0.01). We created a therapeutic scheme to classify our identified target genes into strata based on druggability and approved therapeutics - classifying 41 *novel* targets, 3 *known* targets, and 115 *difficult* targets (of these 69.8% are expressed in the disease relevant cell type from single nucleus experiments). Our *novel* class of genes provides a springboard for new opportunities in drug discovery, development and repurposing in the pre-competitive space. In addition, looking at drug-gene interaction networks, we identify previous trials that may require further follow-up such as Riluzole in AD. We also provide a user-friendly web platform to help users explore potential therapeutic targets for neurodegenerative diseases, decreasing activation energy for the community [https://nih-card-ndd-smr-home-syboky.streamlit.app/].

## Introduction

Currently, there are few approved disease-modifying therapeutics available to those with a neurodegenerative disease (NDD), the most recent being Lecanamab for the treatment of Alzheimer’s disease.^1^ NDDs such as Alzheimer’s disease (AD), Parkinson’s disease (PD), Amyotrophic lateral sclerosis (ALS), Lewy body dementia (LBD), Frontotemporal lobar degeneration (FTLD), and progressive supranuclear palsy (PSP) are diseases caused by progressive nerve cell degeneration that result in a loss of cognition and/or motor function.^2^ The World Health Organization (WHO) expects dementia diagnoses alone to reach 78 million by 2030 and 139 million by 2050. Without disease-modifying therapies, the health, social, and economic impacts of dementia and related NDDs will be catastrophic.^3^ The identification of rational therapeutic targets for NDD will require both the generation of new data and the development and deployment of rapid, open, and transparent tools.

Drugs that are supported by genetic or genomic data frequently outperform those without such evidence in clinical trials. Over two-thirds of the Food and Drug Administration (FDA) approved drugs in 2021 were supported by genetic or genomic evidence.^4^ Therapeutics with genetically supported target mechanisms are twice as likely to pass a trial phase as those without supporting genetic data.^5^ Given the importance of anchoring therapeutic targets to a disease mechanism substantiated by genetic evidence, we developed **omicSynth**: a dynamic, open, and accessible resource that leverages large-scale genetic and genomic data for the identification of therapeutic targets in the neurodegenerative disease space.

The omicSynth resource integrates genetic and genomic data in a Summary-data-based Mendelian Randomization (SMR) framework. The genetic data, primarily in the form of genome-wide association studies (GWAS) from population-scale resources, includes millions of samples across multiple neurodegenerative diseases. Genomic data includes quantitative trait loci (QTL) studies measuring methylation, gene expression, chromatin state, and protein expression. The SMR framework facilitates functional inferences relating disease risk (from GWAS) to the underlying mechanism (from QTL data in relevant cell types). Additionally, we incorporated expression-QTL (eQTL) data from genetically diverse backgrounds into our SMR analyses, addressing the lack of multi-ancestry data in NDD research and allowing for functional inferences regarding differences between ancestral populations.

To add additional context to nominated gene targets, we also investigated single nucleus data from disease-enriched cell types.^6,7^ Many therapeutics mechanistically target inhibition of expression, thus, it is necessary that the target is expressed at baseline in the relevant cell types in order for these treatments to be effective in addressing a particular disease. Using single nucleus expression data, we investigated whether the nominated genes could potentially be affected by inhibitors that are capable of crossing the blood-brain barrier, as in general, genes need to be expressed in order to be susceptible to inhibition from a mechanistic perspective.

We prioritize identified genes as therapeutic targets of interest into three classes based upon known druggability and product market information (Figure 1). *Novel* targets include genes which exhibit significant functional inferences in relevant tissue and cell types in druggable regions of the genome, are not currently targeted by disease-specific therapeutics, and should be prioritized in future repurposing studies. *Known* targets include genes within relevant tissue and cell types that have documented significant functional inferences but are already impacted by a known drug that specifically targets any NDD. *Difficult* target genes are not in regions of the genome currently annotated as druggable. For all novel targets, we searched the corresponding gene regulatory networks to identify companion genes that could also be useful as therapeutic targets. Potential upstream and downstream effects on targeting these genes for therapeutic intervention were provided based on network memberships, and toxicity within these networks was inferred by evaluating liver eQTLs within the network as well as known interacting drugs for each gene of interest. Results can be browsed through the omicSynth resource, which is made available via a free web-based platform, further decreasing activation energy for therapeutic target discovery within the research community [https://nih-card-ndd-smr-home-syboky.streamlit.app/].

**Figure 1:**
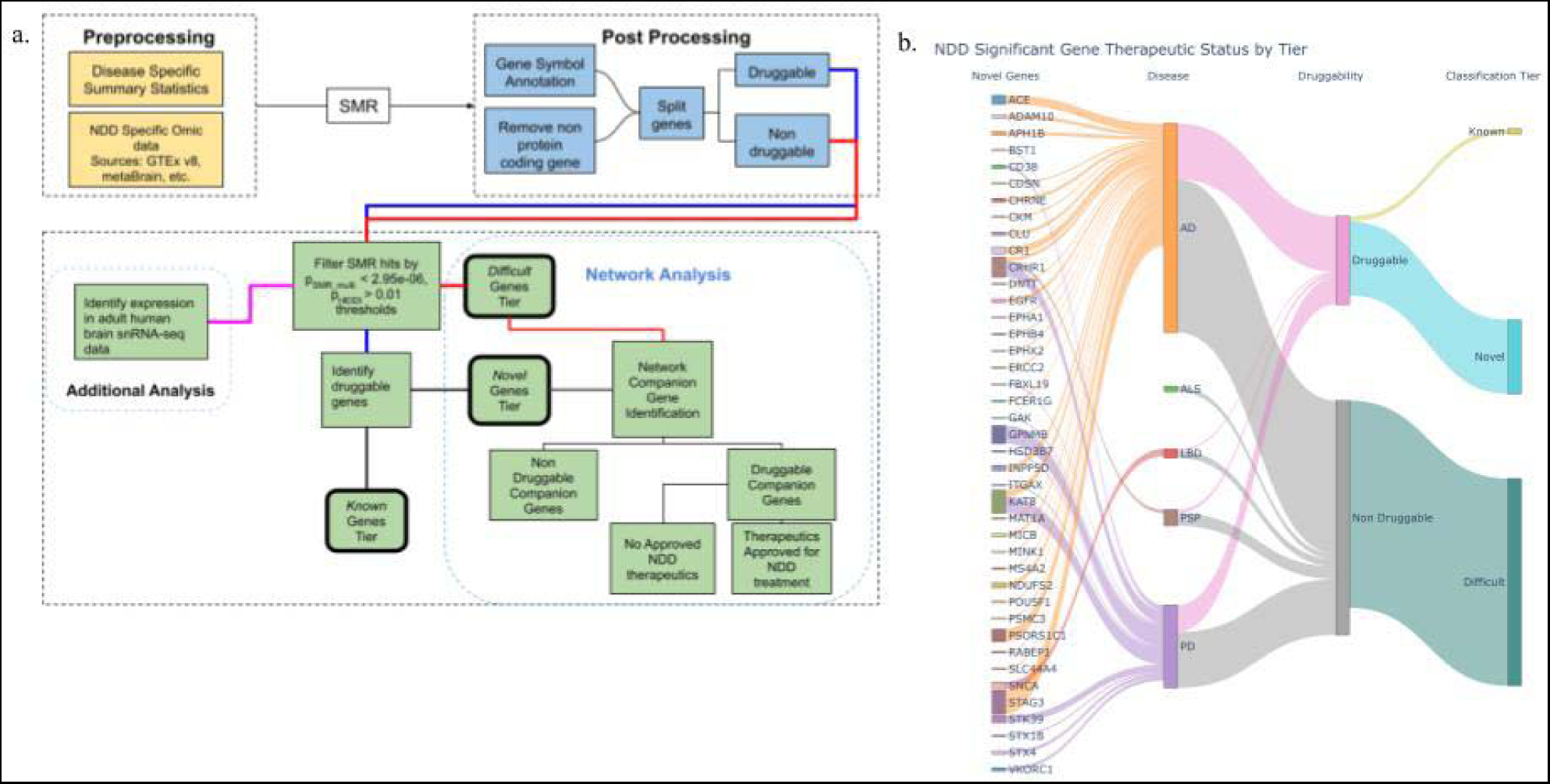
Graphical representation of research workflow and results. **Panel a**: graphical representation of the general workflow used in conducting our analyses. NDD = Neurodegenerative Disease, SMR = Summary-data-based Mendelian Randomization b. Graphical Summary of Results. **Panel b**: Sankey plot depicting the flow of candidate genes into their respective tier. On the left, we highlight *novel* genes, but the remainder of the plot visualizes all 159 candidate genes regardless of the final classification tier.

## Results

### Overview

We identified 540 candidate gene-level SMR associations (159 unique gene targets) across six NDD and 186 tissue-omic pairings with a stringent disease-level multiple test correction threshold (p_SMR_multi_ < 2.95×10^−6^ and p_HEIDI_ > 0.01; **Supplementary Tables S1, S2, S4**). On a per-disease basis we identified 317 total significant associations: 116 unique gene targets for AD, 4 significant associations across 3 unique gene targets for ALS, 13 significant associations across 5 unique genes for LBD, 184 significant associations across 46 unique gene targets for PD, and 22 significant associations across 9 unique gene targets for PSP. FTLD had no significant associations at our corrected p-value threshold. No NDD showed significant pQTL or caQTL associations. Of the 159 unique genes across all diseases, 69.8% (111 genes) were expressed in tissues enriched with disease-associated gene expression and 11.9% (19 genes) showed colocalization with brain tissue eQTLs with posterior probability (PP) > 0.90.^7^ Colocalized genes from this complementary analysis are also bolded in **Table 1** and **Table 2**.

**Table 1:** Candidate genes for multiple neurodegenerative diseases. This table shows genes with functional inferences passing multiple test corrections for multiple neurodegenerative diseases. We provide details for all the omics and diseases in which a given gene has significant associations. Bolded genes indicate colocalized genes.

Tables with Legends
| Gene | Diseases | Omics |
| --- | --- | --- |
| ARL17B | AD, PD | Cerebellum eQTL, Cortex eQTL, Spinalcord eQTL |
| KAT8 | AD, PD | Cerebellum eQTL, Whole Brain meta-analysis mQTL, Cerebellar Hemisphere eQTL, Cortex eQTL, Tibial Nerve eQTL, Skeletal Muscle eQTL, Hypothalamus eQTL, Whole Brain eQTL, Cerebellum eQTL, Spinalcord eQTL |
| LRRC37A2 | AD, PD | Hippocampus eQTL, Cortex eQTL, Frontal Cortex BA9 eQTL, Prefrontal Cortex eQTL, Caudate Basal Ganglia eQTL, Skeletal Muscle eQTL, Multi Ancestry, Whole Brain Meta-analysis eQTL, Hypothalamus eQTL, Liver eQTL, Anterior Cingulate Cortex BA24 eQTL, Putamen Basal Ganglia eQTL, Amygdala eQTL, Whole Brain eQTL, Cerebellum eQTL, Nucleus Accumbens eQTL, Basal Ganglia eQTL, Spinalcord eQTL, Hippocampus eQTL, Substantia nigra eQTL |
| KANSL1 | AD, PD, PSP | Whole Brain meta-analysis mQTL, Whole Blood mQTL, Cortex eQTL, Multi Ancestry Whole Brain Meta-analysis eQTL, Spinalcord eQTL, Anterior Cingulate Cortex BA24 eQTL |
| ARL17A | AD, PD, PSP | Spinalcord eQTL, Amygdala eQTL, Multi Ancestry Whole Brain Meta-analysis eQTL, Hypothalamus eQTL, Hippocampus eQTL, Cerebellar Hemisphere eQTL, Cortex eQTL, Caudate Basal Ganglia eQTL, Anterior Cingulate Cortex BA24 eQTL, Putamen Basal Ganglia eQTL, Cerebellum eQTL, Nucleus Accumbens Basal Ganglia |
| PRSS36 | AD, PD | Whole Brain meta-analysis mQTL, Cortex eQTL, Cerebellar Hemisphere eQTL, Multi Ancestry Whole Brain Meta-analysis eQTL, Whole Brain eQTL |
| MAPT | AD, PD, PSP | Whole Brain meta-analysis mQTL, Whole Blood mQTL |
| IDUA | LBD, PD | Whole Brain meta-analysis mQTL, Whole Blood mQTL, Whole Blood eQTL(eQTLgen) |
| TMEM175 | LBD, PD | Whole Blood mQTL |
| ARHGAP27 | AD, PD, PSP | Whole Blood mQTL, Whole Blood eQTL (eQTLgen), Multi Ancestry Whole Brain Meta-analysis eQTL, Caudate Basal Ganglia eQTL, Nucleus Accumbens Basal Ganglia |
| CRHR1 | AD, PD, PSP | Whole Brain meta-analysis mQTL, Whole Blood mQTL, Cortex eQTL, Skeletal Muscle eQTL |
| FMNL1 | AD, PSP | Multi Ancestry Whole Brain Meta-analysis eQTL, Whole Blood mQTL |
| PLEKHM1 | PD, PSP | Cortex eQTL, Frontal Cortex BA9 eQTL, Prefrontal Cortex eQTL, Caudate Basal Ganglia eQTL, Skeletal Muscle eQTL, Anterior Cingulate Cortex BA24 eQTL, Putamen Basal Ganglia eQTL, Whole Brain eQTL |
| WNT3 | AD, PD | Cortex eQTL metaBrain, Skeletal Muscle eQTL, Tibial Nerve eQTL |
| SPPL2C | AD, PD | Cerebellum eQTL, Prefrontal Cortex eQTL |

**Table 2:**
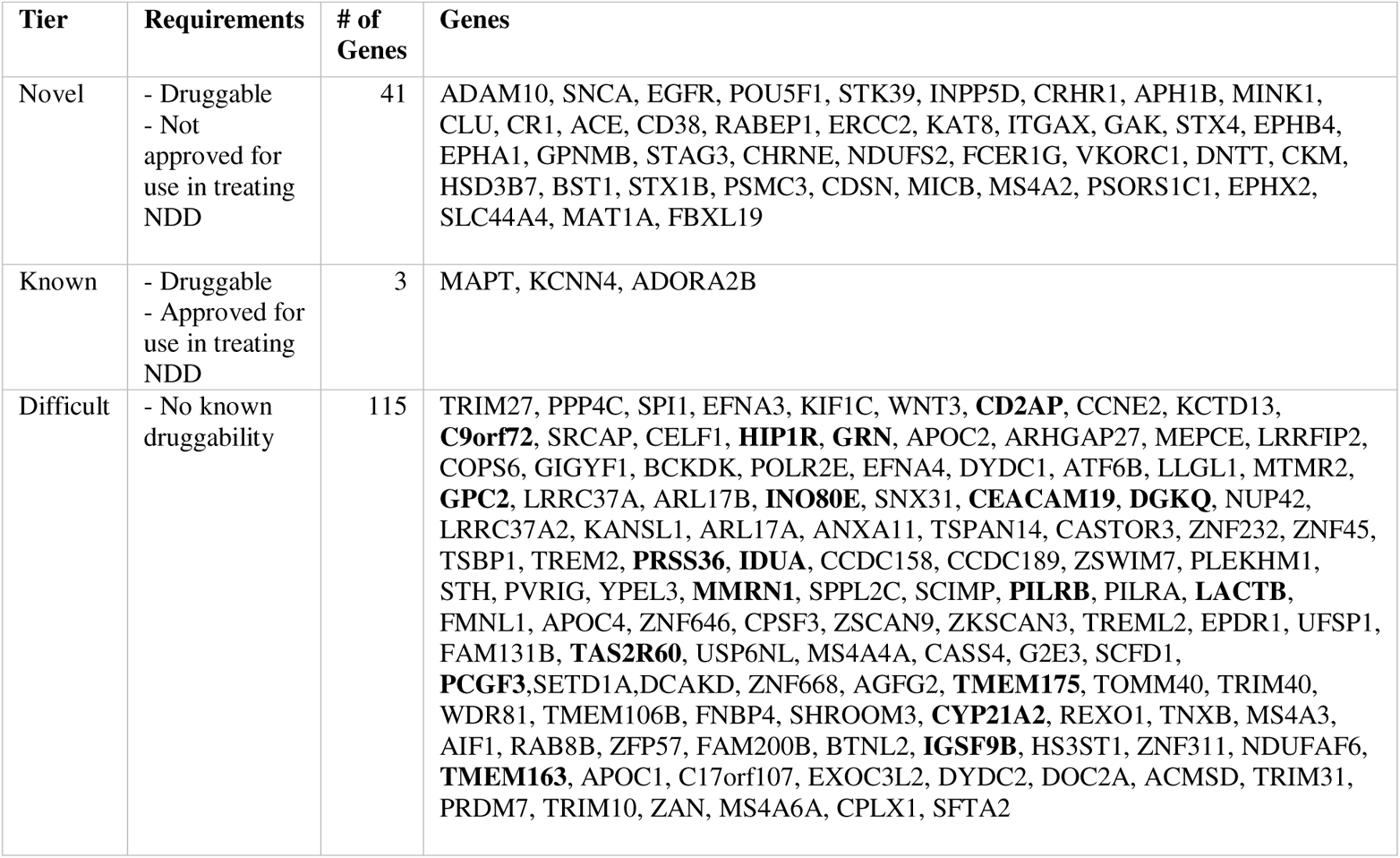
Therapeutic Classification Scheme by Tier. Table providing information on the three classifications tiers in our therapeutic classification scheme including requirements for each tier. Bolded genes indicate colocalized genes.

### SMR analysis identifies 15 common genes significant across multiple NDD

Using SMR, we identified 15 unique genes across 182 associations to be significant in two or more NDDs at a stringent significance threshold (p_SMR_multi_ < 2.95×10^−6^ and p_HEIDI_ > 0.01; **Table 1**, **Supplementary Tables S2-S4**). Of the identified genes, five genes (*MAPT, CRHR1, KANSL1, ARL17A,* and *ARHGAP27*) were found to be significant across 97 tested associations and three NDDs (AD, PD, and PSP). *MAPT* and *CRHR1* were found to be largely significant in mQTL omics with *MAPT* significant in whole blood and brain mQTL data for all previously mentioned NDDs and *CRHR1* found to be significant in whole blood mQTL data for all three NDDs (Supplementary Tables S3 – S5). Additionally, only *MAPT* and *CRHR1* are annotated as druggable in multiple drug data sources as of the writing of this manuscript. All genes, except for *ARL17A*, had multiple significant associations in both brain and blood mQTL tissues. *ARHGAP27* and *KANSL1* had significant associations replicated in the multi-ancestry eQTL data (African, American, East Asian, European, and South Asian ancestries), suggesting a potential generalizability of these targets across different populations.

We identified 10 unique genes across 85 significant associations in any two NDD (p_SMR_multi_ < 2.95×10^−6^ and p_HEIDI_ > 0.01). One of the identified genes is considered therapeutic and the remaining nine are non-therapeutic. *KAT8*, *ARL17B, PRSS36, LRRC37A2, WNT3,* and *SPPL2C* were all found to be significant in both AD and PD, *IDUA* and *TMEM175* were found to be significant in LBD and PD, *PLEKHM1* was significant in *PD* and *PSP*, and *FMNL1* was significant in AD and PSP (**Supplementary Tables S3-S5**). *KAT8,* the only therapeutic gene, showed significant associations with decreased expression in brain tissue and blood, as well as a significant association with increased methylation. Of the remaining nine genes, *IDUA, FMNL1, PRSS36,* and *TMEM175* had significant associations in mQTL sources. Additionally, *FMNL1, LRRC37A2,* and *PRSS36* had significant associations replicated in the multi ancestry eQTL data (p_SMR_multi_ < 2.95×10^−6^; **Supplementary Table S7**).

### Drug Target Discovery using significant genes identifies 41 novel gene targets for follow-up study

Using the approach previously outlined in our introduction and methods for drug target gene nomination, we categorized 159 gene hits into one of three tiers (**Table 2**). In our first tier, *novel* genes, we nominated 41 gene targets. SMR results for the *novel* genes are listed in **Supplementary Table S8**. Genes are categorized as *novel* if they are in druggable regions of the genome that can be targeted by common molecular methods and currently have no FDA-approved treatment for any NDD as identified by current literature, knowledge base, and drug databases. Our second tier, *known* genes, had three gene targets identified - *MAPT, KCNN4,* and *ADORA2B*. *Known* genes are genes that have at least one FDA-approved therapeutic for treatment of an NDD (**Supplementary Table S9**). The 3 nominated *known* gene targets are targeted by four therapeutics - Apomorphine, Carbidopa, Istradefylline, and Riluzole. Currently these therapeutics are used for the treatment of PD symptoms (Apomorphine, Carbidopa, Istradefylline) and prolonged survival for ALS (Riluzole). In our last and largest tier, *difficult genes*, we identified 115 gene targets with no currently known therapeutics that target these genes and no known druggability. A total of 52 of the identified *difficult* genes exhibited at least two significant associations, with *LRRC37A2* having the maximum number of significant associations at 25 associations across AD and PD.

### Network analysis provides insight into druggable companion genes to non-druggable genes of interest

We further implemented a gene network analysis for our *novel* and *difficult* tier candidates to identify potential proxy gene targets within each nominated genes Signor curated network. In the *novel* tier gene, we identified 87 companion genes of which 58 are considered potentially druggable (**Supplementary Table S9**). Of the 58 druggable companion genes, 30 were found to be targeted by a *known* drug and a further five are targeted by therapeutics approved for treatment of AD. The five companion genes with AD targeted therapeutics are *NCSTN, MAPK14, PSEN1, PSEN2,* and *PSENEN*. Genes *NCSTN, PSEN1, PSEN2,* and *PSENEN* are all targeted by Tarenflurbil, Semagacestat, and Avagacestat, while *MAPK14* is targeted by Neflamapimod, an oral p38 alpha kinase inhibitor that the FDA approved for use in the treatment of AD and LBD (**Figure 2**). Further analysis of *difficult* genes co-expression networks identified 27 genes with 65 curated companion genes (**Supplementary Tables S10-S12**, **Supplementary Figure S1**). Of the 65 identified companion genes for the *difficult* target tier, 34 were found to be druggable with 18 having *known* drugs. *MAPK14* was the only companion gene to have a therapeutic approved to treat an NDD in the *difficult* target tier. *MAPK14* was identified as a companion gene to the *difficult* gene *TRIM27* (**Figure 2**).

**Figure 2:**
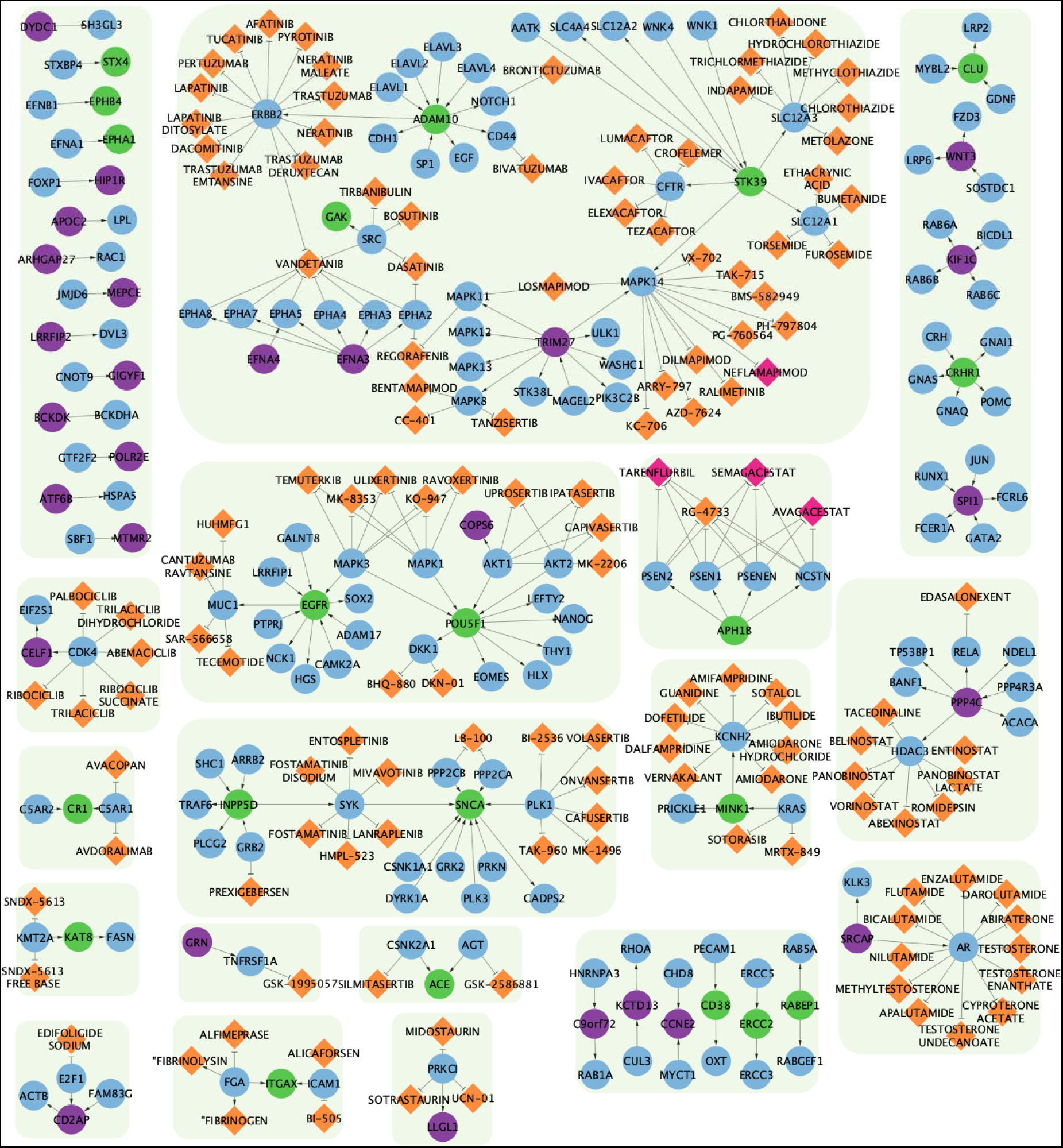
Network Visualization of *Novel* and *Difficult* genes companion genes and drugs. Graph network visualization of both *novel* (green nodes) and *difficult* (purple nodes) genes and their SIGNOR curated partners (blue nodes). Drugs that interact with companion genes are denoted by orange-colored diamonds while FDA-approved drugs for use in NDD treatment are colored pink. Connecting arrows indicate the direction from regulator gene to target gene.

### Multi-ancestry analyses reveal opposing gene expression patterns in significant disease risk loci between Non-European and European ancestries

To investigate our findings in more diverse data, we also performed SMR with multi-ancestry eQTL data, aiming to replicate previous significant results (p_SMR_multi_ < 2.95×10^−6^ and p_HEIDI_ > 0.01) and nominate targets that show evidence towards generalizability across multiple ancestry groups. After multiple test correction, we significantly replicated 11 total associations corresponding to 9 genes that were nominated in our initial analysis: *ARHGAP27*, *ARL17A*, *GPNMB*, *KANSL1*, *LRRC37A2*, *PILRA*, *PILRB*, *PRSS36*, and *ZNF232* (see **Supplementary Table S7**).

We then categorized these nine replicated genes based on their druggable status. One of the replicated hits, *GPNMB*, is classified in our *novel* gene tier, meaning that it is druggable and has no known treatment approved for treatment of NDDs, which suggests this gene may be an interesting but also potentially generalizable target across ancestries. The remaining eight replicated genes were in our *difficult* tier, meaning they are currently considered non-druggable.

### Nominated genes of interest found to be expressed in disease relevant adult human brain snRNA-seq expression data

To provide additional evidence for biological relevance of our nominated targets, we investigated whether the 159 significant genes nominated through SMR (P_SMR_multi_ < 2.95×10^−6^ & P_HEIDI_ > 0.01) were expressed in relevant cell types from adult human brain snRNA-seq data.

We tested the 159 genes found to be significant against adult human snRNA-seq data to identify expression in varying brain single cell types (P_SMR_multi_ < 2.95×10^−6^ & P_HEIDI_ > 0.01). We calculated the mean and median expression percentile rank (EPR) for each gene across cells corresponding to each of the 31 tested cell types. To ease interpretation, we additionally binned the mean EPR values into three expression categories: *off, low,* and *high* based on the mean EPR value for each gene-cell type combination. Using our binned mean EPR values we found that 40 genes had all exclusively *off* EPR values while 16 genes had exclusively all *low* EPR values across all 31 tested cell types (**Figure 3, Supplementary Tables S13-S14**). We identified 11 genes (*KANSL1, LRRFIP2, CELF1, MAPT, STK39, RABEP1, SCFD1, CLU, SNCA, TMEM163,* and *SHROOM3*) that had at least one gene highly expressed in any single cell type with *KANSL1* having the most *high* EPR values across 15 cell types (**Figure 3**, **Supplementary Tables S13-S14**). Detailed breakdowns of both mean and median EPR values are provided in **Supplementary Figures S2-S4** and **Supplementary Tables S13-S14**. Cell types Hippocampal CA4 and Deep layer intratelencephalic had the most genes with *high* EPR values and Hippocampal CA4 had the least number of genes with *off* EPR values (n_hig_h= 5, n_low_=89, n_off_=65) The Vascular cell type had the highest number of genes with *off* values (n_off_=123).

**Figure 3:**
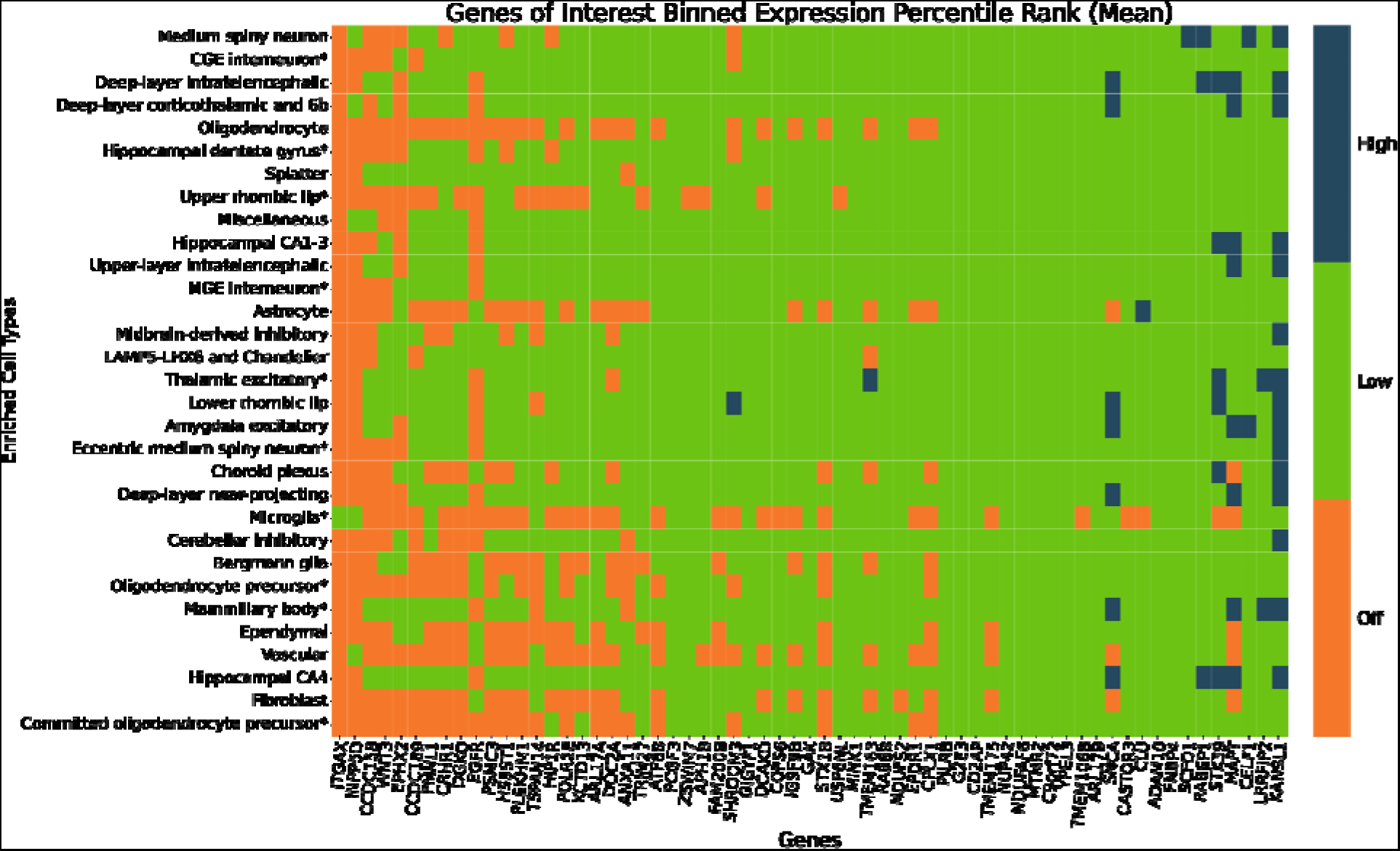
scRNA-seq expression for significant genes (p_SMR_multi_ < 2.95×10^−6^ and p_HEIDI_ > 0.01). Expression of a given gene within each cell type is categorized as being highly expressed (dark blue, top 10% of all genes), intermediately expressed (green, middle 80%) or undetected (orange, bottom 10%). Genes that had *off* mean expression percentile rank values across all tested cell types were excluded from the heatmap. Cell types found to be enriched with AD and/or PD-relevant genes are marked by an asterisk.

We also investigated the nominated genes in disease relevant cell types - Bergmann glia, CGE interneuron, Committed oligodendrocyte precursor, Deep layer intratelencephalic, Eccentric medium spiny neuron, Hippocampal CA1 3, Hippocampal dentate gyrus, LAMP5 LHX6 and Chandelier, MGE interneuron, Mammillary body, Microglia, Midbrain derived inhibitory, Oligodendrocyte precursor, Thalamic excitatory, Upper layer intratelencephalic, Upper rhombic lip - identified in Alvarado et al to be significantly enriched with AD and PD-relevant genes.^7^ Significant genes from our SMR analysis were highly expressed in three of these disease relevant cell types, Thalamic excitatory, Eccentric medium spiny neuron, and Mamillary body. *KANSL1* had high expression in all three listed cell types (**Figure 3**). Of the 159 tested genes, 20 had all *low* EPR values in the disease relevant cell types. The *thalamic excitatory* cell type had the least number of *off* EPR values for genes *INPP5D*, *ITGAX*, *EGFR*, and *DOC2A*.

## Discussion

As the global population continues to age, the threat posed by NDDs presents a behemoth and multifaceted challenge. Our research aims to address the challenge of treating NDDs by identifying therapeutic targets anchored in genetic data - a proven strategy in therapeutic development. Implementation of this strategy has been impeded by the small sample sizes and the dispersed nature of genetic and disease-related data, such as proteomics and transcriptomics. Here, we attempted to address this need by creating and implementing an open-source framework t identify druggable targets across varied NDDs.

In our targeted analyses, we were unable to identify any potentially functionally important genes that were significant across all six tested NDDs. While NDDs share prominent hallmarks, such as cell death, inflammation, and pathological protein aggregation, the role that each hallmark and its associated biological processes plays in the pathogenesis of each NDD differs, creating a spectrum.^8,9^ We identified *MAPT*, *CRHR1*, *KANSL1*, *ARL17A*, and *ARHGAP27* to be significant in multiple different omics for AD, PD, and PSP (**Supplementary Tables S3-S5**). *MAPT* was found to have significant associations with primarily increased expression for AD, PD, and PSP across eQTL and mQTL omic data, as supported by previous research.^10–12^ The *MAPT* locus, 17q21, contains genes *CRHR1*, *KANSL1*, *ARL17A*, and *ARHGAP27*, and mutations in this locus have been previously associated with both PD and PSP.^13^ Previous evidence of significant association of this locus in AD is more fragmented and sparse. The 17q21 locus which includes genes *KANSL1* and *MAPT* has been previously implicated in AD.^14^ *ARL17A* has been reported to harbor eQTL SNPs implicated in both brain and blood tissues in relation to AD.^15^ *CRHR1*’s role in stress response has been hypothesized to exacerbate AD pathologies given its abundance in the brain including areas implicated in learning and memory.^16^ Lastly, evidence of *ARHGAP27*’s significance in AD includes associations between complex traits such as cognitive functioning, reaction time, and cortical structure phenotypes.^17,18^

A deep dive into *KANSL1* highlights its role in autophagy pathways. *KANSL1* is a core member of the nonspecific lethal (NSL) complex that binds to MOF (also known as *KAT8*) which is necessary for the acetylation of histone H4 lysine 16 acetylation (H4K16ac).^19,20^ Some studies have associated elevated expression levels of *KANSL1* with over promoted autophagic activity resulting in cell death and cytotoxicity from autophagosome accumulation;, although further research is required to understand this mechanism.^21^ Additional research into the role of autophagy and lysosomal pathways in NDD have indicated that altered autophagy function results in the inability to clear out protein aggregates resulting in cell death and potentially contributing to disease pathogenesis and neurodegeneration. ^20,22–24^ Our results are consistent with previous research linking increased expression of *KANSL1* with neurodegenerative effects. When assessing associations with AD, PD, and PSP, *KANSL1* is associated with an increased expression in brain mQTLs, three different brain eQTLs (psychEncode, multi ancestry, and anterior cingulate cortex), and spinal cord eQTLs. The consistent significance of *KANSL1* and most of our gene hits in mQTL omics highlights the influence of DNA methylation for NDD pathogenesis and progression.

We identified 10 genes as significant in two diseases. The nominated genes do not share any explicit relationships but are common in their importance for varying biological processes and cellular functions such as cell proliferation and differentiation, degradation of transmembrane proteins, calcium homeostasis, and autophagy regulation [50-54]. ^25–29^ Six of our nominated genes, *ARL17B, KAT8, LRRC37A2, PRSS36, SPPL2C,* and *WNT3*, are associated with both AD and PD. Given the significantly larger sample sizes and increased power of the two diseases in GWAS summary statistics, we did not find this unexpected. LBD and PD share two genes, *IDUA* and *TMEM175,* while AD and PSP share *FMNL1* and PD and PSP share *PLEKHM1* (Supplementary Tables S3, S4, S6). In general, the bulk of the gene hits were found to be significant in mQTL data for both brain and blood tissues (n_whole brain_= 4; n_whole blood_= 4) followed by cortex eQTLs (n_cortex metaBrain_ = 6, n_cortex GTEx_ = 3, n_Frontal Cortex BA9_ = 2, n_prefrontal cortex_ = 3).

The only gene found in two diseases, AD and PD, that could be targeted therapeutically was *KAT8,* which we previously mentioned in the context of the *KANSL1* gene. In literature, *KAT8* (Lysine Acetyltransferase 8) is identified as a protein-coding gene that plays a vital role in the NSL complex for acetylation of H4K16ac.^24^ Scientific observation has identified the consequences of autophagic dysfunction in NDD to include impaired neuronal function, neuronal death, and neuron loss. In opposition to the expression pattern of *KANSL1*, decreased expression of *KAT8* is associated with deacetylation of H4K16ac in AD patients, while an overexpression of KAT8 has been linked to increased expression levels of neuroprotective soluble amyloid precursor protein (sAPP)α and β-secretase (BACE)2 and decreased levels of sAPPβ and BACE1.^30^ In our results, we found blood mQTLs for AD and brain mQTLs for AD and PD to be associated with increased expression of *KAT8*; this is in contrast to gene expression in some of the same tissues, such as blood and brain mQTLs, for *KANSL1*. The associated increased expression of *KAT8* in our results suggest that an increase in expression may be correlated with excess autophagy resulting in cell death, which is a hallmark symptom of all three NDD (AD, PD, and LBD).^31,32^ There currently does not exist any FDA approved therapeutics that target *KAT8* in NDD. However, compound MG149, a histone acetyltransferase inhibitor, has been found to reduce proinflammatory genes via inhibition of MYST type histone acetyltransferase KAT8.^33^ MG149 has also been found to be effective in restoring impaired autophagic flux via the inhibition of histone acetylation of H4K16ac in cases of ischemic stroke and inflammatory diseases.^23,34^ Further research into the application of MG149 could result in a novel treatment targeting the characteristic accumulation of toxic proteins in NDD.

FTLD was the only tested disease that did not have any suggestive targets at our test correction threshold. This may be because the FTLD GWAS had the smallest sample size out of all the diseases tested, and results will likely improve as larger FTLD GWAS are conducted. As there were no significant results for FTLD after correction, we decided to investigate potential pleiotropic relationships between FTLD and the other NDDs. To do this, we looked for FTLD associations at a less-stringent P value threshold (p_SMR_multi_ < 0.05) only in the 254 unique candidate genes passing our original threshold of p_SMR_multi_ < 2.95×10^−6^, a process detailed by Baird et al.^35^ This resulted in 124 FTLD hits made up of 31 unique genes that have a potential pleiotropic relationship between FTLD and another NDD. Of those 31, 12 were classified as druggable through our sources (*STX4*, *STX1B*, *VKORC1*, *POU5F1*, *HSD3B7*, *PSORS1C1*, *SLC44A4*, *CD38*, *EPHX2*, *FBXL19*, *CLU*, *CDSN*). All 12 fall into the novel tier of drug targets, representing potential avenues for drug repurposing for FTLD.

Our creation of a drug target classification scheme is an attempt to inform drug discovery and repurposing from genes considered significant with evidence of causative roles in NDD. Further inspection of our 41 *novel* genes provides multiple insights into the genes that compose the tier. Many genes that compose our novel tier have therapeutics used in the treatment of multiple types of cancers and tumors. Fourteen of our novel genes have therapeutics approved for use in the treatment of cancer (MONDO_0004992). Other common approved indications for therapeutics that target our novel genes include, but are not limited to, neoplasm (EFO_0000616), hypertension (EFO_0000537), and cardiovascular disease (EFO_0000319). *GPNMB,* which is of particular interest due to support for its role in PD, falls into this grouping of 14 genes. Similar to its role in cancer and tumor growth, our results highlight *GPNMB*’s pattern of increased expression as shown in brain related PD eQTLs. We were able to find replication of increased GPNMB expression in brain related tissues in Li et al, Ortiz et al, and Nalls et al. ^36–38^ Glembatumumab Vedotin is one of the therapeutics that targets *GPNMB* where its primary mechanism of action (MOA) is Tubulin inhibition.^39^ Consequently, Glembatumumab Vedotin’s inhibitory MOA could be repurposed for use in PD treatment for suppression of inflammation given the recognized role of inflammatory response/neuroinflammation in PD onset and progression.^40,41^ However, any treatment developed targeting *GPNMB*, would most likely be limited in treating people of European ancestries due to the gene’s importance and role compared to non-European ancestries - further increasing inequality.

Our largest and most uncertain classification tier contains 121 *difficult* genes. Despite not having any currently known therapeutics, this classification tier could lead to the development of NDD targeted therapeutics or the repurposing of existing ones. Our approach for these genes focused on analyzing well curated networks centered on each *difficult* gene to identify any partner genes with existing therapeutic drugs. This approach provides us context into any biological pathways and processes that may be affected by a targeted treatment which could help eliminate the time and resources spent on developing and researching ineffective therapies.

The smallest tier, *known* genes, is composed of the three genes targeted by NDD targeted therapeutics. Apomorphine, Carbidopa, and Istradefylline are indicated for use in treatment of PD. Riluzole is indicated for the treatment of ALS but has undergone phase 2 clinical trials for use in treatment of AD. The results in clinical trials for use of Riluzole in AD treatment were promising with cerebral glucose metabolism, an AD biomarker, preserved in patients receiving riluzole compared to those in the placebo group [66].^42^ The researchers conducting the study suggested a more powerful and longer study, but no follow up studies have yet been initiated. Our results support the continued follow up of Riluzole clinical trials.

Focusing on genes we flagged as putatively associated with risk across multiple diseases, 13 of 15 were noted as being at least moderately expressed in cell types of interest (those with enriched expression for GWAS risk signatures) from single nucleus sequencing. Positive beta coefficients at these genes from the SMR analysis suggest that if an expression effect was inhibited, it could be possible to reduce disease risk. Two of these genes, *KANSL1*, and *MAPT*, showed significant positive associations (defined as a gene with positive beta values in more than 50% of its significant SMR associations) between risk and expression in our SMR analyses, providing a contextual insight for future follow-up.

Genes such as *GPNMB* had different expression patterns in European and non-European ancestries. For example, *GPNMB* had decreased associated expression in multi ancestry eQTLs but an increased associated expression in all other tested eQTLs. Previous research in certain Asian populations has found no significant association between *GPNMB* and PD [67,68].^43,44^ Rizig and colleagues, conducting the largest PD GWAS in the African and African admixed populations in ~200,000 individuals, of which 1,488 are cases, report the following per SNP in *GPNMB*: rs858275, P=0.1250, beta=-0.0824, indicating no association in African/African admixed ancestries. Our multi ancestry data reports the same direction of expression in GPNMB SNP rs858275, P=1.080397×10^−8^, beta=-0.107745 in PD. Interestingly, the reported direction of expression in our multi ancestry data and Rizig and colleagues’ data is in contrast to the direction of expression reported for European ancestries, in addition to indicating no significant associations (**Supplementary Table S13**, unpublished data).^45^

The limitations we encountered in our research included limited GWAS data for diseases excluding AD and PD, limited non-eQTL omic data, as well as limited multi ancestry omic data and reference panels. In general, the availability of publicly and freely available omic data is consistently increasing. As new data is published, we intend to conduct updates and incorporate new omic types into our analysis such as more pQTL, single cell QTLs, and splicing quantitative trait loci (sQTLs). The incorporation of additional multi omic data should provide new and novel insights into the complex underpinnings of NDD.

The limitation we feel presents the most barriers is that of limited multi ancestry data. The state of diversity in the NDD research space has historically been Eurocentric which remains the case in this study due to the limited availability of genetic data from non-European participants. One of the distinguishing aspects of this study is the inclusion of multi ancestry eQTL data in the search for generalizable drug targets. This is particularly important in an era where precision medicine and machine learning can introduce inherent bias when the only reference data is from European ancestry populations. We identified common hits which are consistent with current understanding that there are NDD risk loci that are shared across genetic ancestries while providing insight on which gene loci and differences in expression may play a role in NDD development and treatment in non-Europeans. It is worth noting that while replication was limited at our stringent significance threshold, we were able to make some interesting observations. While we made attempts to include a limited set of multi ancestry in the future we would like to be able to include more multi ancestry disease GWAS and omic data to make more meaningful insights. We look forward to the increasing availability of non-European data with the creation of data sources such All of Us, an NIH research program focusing on inclusion of health data of marginalized populations in the United States.^46^

This report is a description of the foundation for a community driven resource to identify and investigate future genetically derived drug targets in an open-source context. Ultimately, we are working on creating a network tool that incorporates multi-omic data, disease GWAS summary statistics, drug data, and other relevant data types to ease research such as this study, eliminating barriers to drug discovery and drug repurposing and potentially enabling precision medicine in the NDD space. Using multi omics integration methods, deep learning techniques, and most importantly, community input to better parse and interpret the data presented by the platform we aim to make our community resource a robust tool for NDD research.

## Data Availability

Omic data and GWAS summary statistics which we have expanded on in our Methods section are freely and publicly available. An extensive listing of availability, as well as direct links to the data sources, can be located here (https://nih-card-ndd-smr-home-syboky.streamlit.app/About). All code can be found on the NIH CARD github linked here (https://github.com/NIH-CARD/NDD_SMR). All SMR associations and annotations are available to browse using an interactive web application (https://nih-card-ndd-smr-home-syboky.streamlit.app/).

https://nih-card-ndd-smr-home-syboky.streamlit.app/About

https://github.com/NIH-CARD/NDD_SMR

https://nih-card-ndd-smr-home-syboky.streamlit.app/

## Acknowledgements

We would like to thank all the subjects who donated their time and biological samples to the studies that are reflected in these analyses. This research was supported in part by the Intramural Research Program of the National Institutes of Health (National Institute on Aging and National Institute of Neurological Disorders and Stroke; project numbers: 1ZIANS003154, Z01-AG000949-02, ZIAAG000534). This study used the high-performance computational capabilities of the Biowulf Linux cluster at the National Institutes of Health (http://hpc.nih.gov).

## Author Contributions

M.A.N., C.X.A., D.V., H.L.L., F.F., and H.I conceived and planned the main conceptual ideas. M.A.N., H.L.L., and F.F supervised the project. D.V., C.X.A, and M.A.N. performed all initial data collection, wrangling and computations. C.X.A. and C.A.W. processed and updated all experimental data, performed the analysis, and designed the figures. H.L.L., M.B.M., and M.A.N. aided in interpreting analyses results. C.X.A, M.B.M., M.A.N., and C.A.W. contributed to the drafting of the manuscript and preparation for publication. A.S., K.L., M.J.K., and S.B.C. consulted and provided critical feedback to the manuscript. All authors revised the manuscript.

## Declaration of Interests

CXA, DV, KL, HLL, FF, and MAN declare that they are consultants employed by Data Tecnica International, whose participation in this is part of a consulting agreement between the US National Institutes of Health and said company. MAN also an advisor to Neuron23 Inc and Character Biosciences.

## STAR Methods

### Key Resources Table

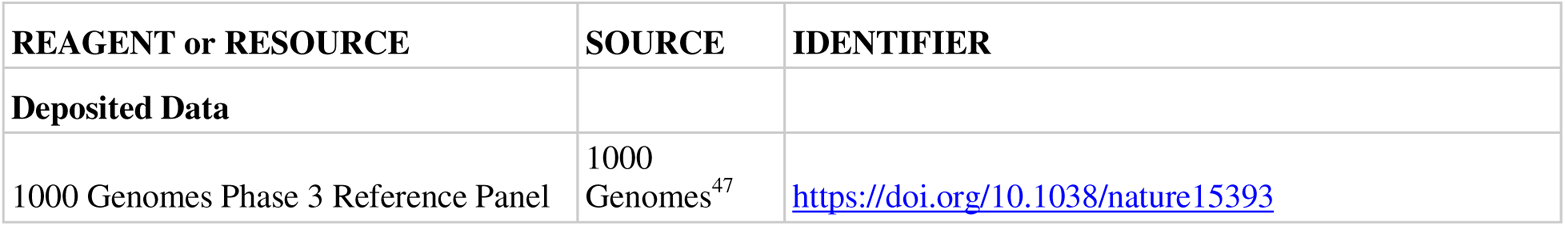
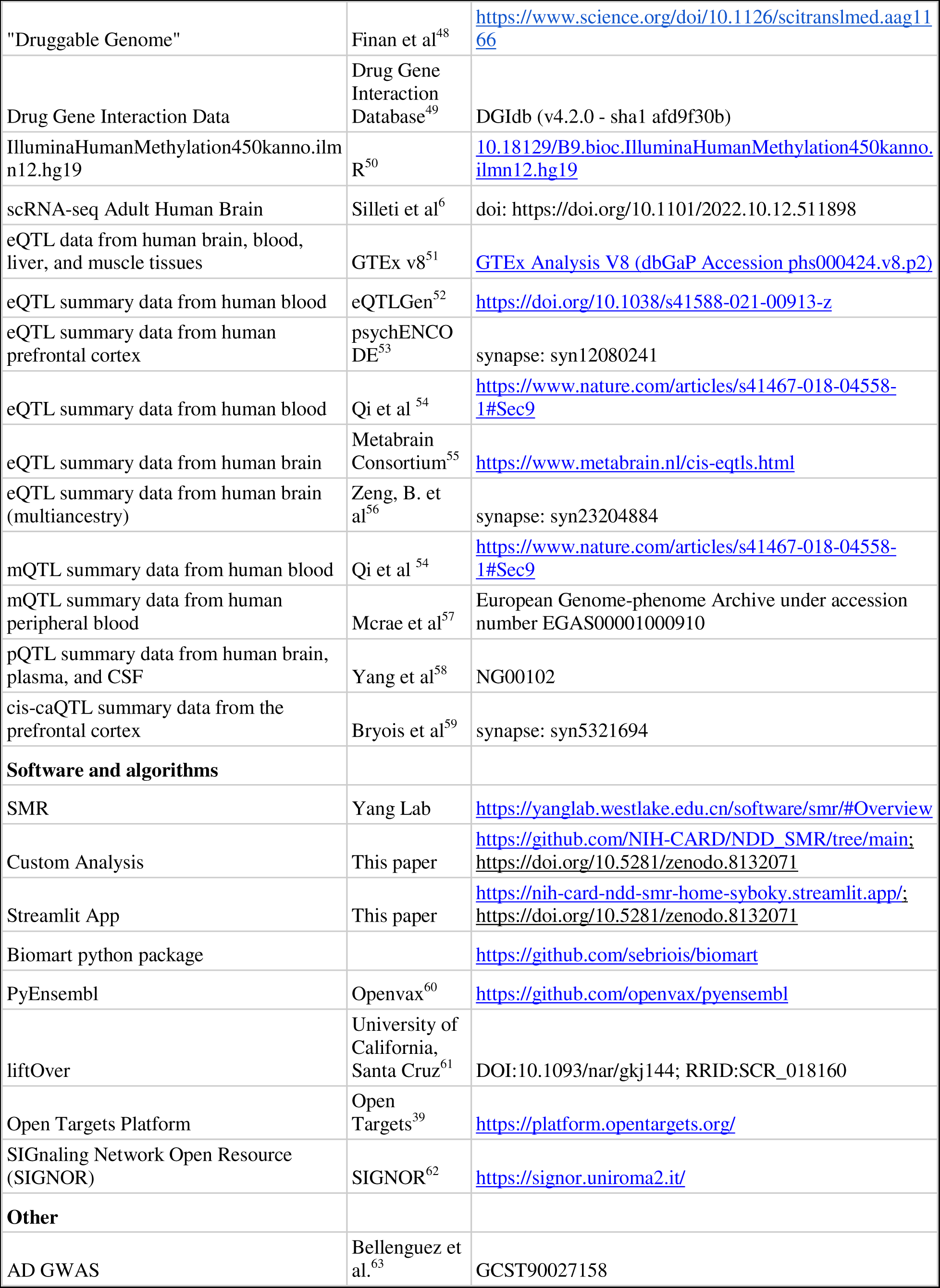
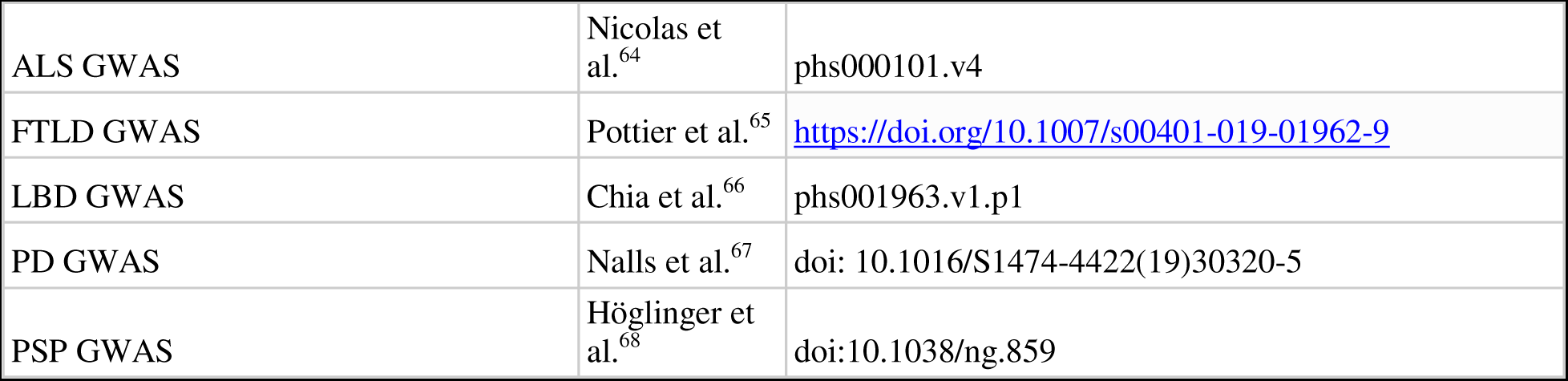

### Resource Availability

#### Lead Contact

Further information and requests for resources and data should be directed to and will be fulfilled by the lead contact, Chelsea Alvarado.

#### Materials availability

This study did not generate new unique reagents.

#### Data and code availability

- This paper analyzes existing, publicly available data. These accession numbers for the datasets are listed in the key resources table.
- All original code has been deposited at GitHub, deposited on Zenodo and is publicly available as of the date of publication. GitHub URL and Zenodo DOI are listed in the key resources table.
- Any additional information reported in this paper is available from the lead contact upon request.

### Method Details

#### Datasets

Genome-Wide Association Studies (GWAS) summary statistics for each of the six NDD highlighted in our study were used to obtain SNPs that served as instrumental variables in the Mendelian randomization pipeline. GWAS used are the latest and/or largest for each corresponding disease: Bellenguez et al. (2022) for AD (n = 788,989); Chia et al. (2021) for LBD (n = 7,372); Höglinger et al. (2011) for PSP (n = 4,361); Nalls et al. (2019) for PD (n = 1,456,306); Nicolas et al. (2018) for ALS (n = 80,610); and Pottier et al. (2019) for FTLD (n = 1,355)^63–68^.All GWAS summary statistics were lifted over, as needed, to hg19 (GRCh37) using University of California, Santa Cruz’s liftOver command line tool.^61^

#### x-QTL Summary Statistics

Expression quantitative trait loci (eQTL), protein quantitative trait loci (pQTL), chromatin quantitative trait loci (caQTL) and methylation quantitative trait loci (mQTL) were used as the exposure variables in the Mendelian randomization (MR) analyses. eQTLs are genetic loci that explain the variation in mRNA expression levels. Cis-eQTL, eQTLs that act on local genes, data makes up most all x-QTL data used for our study due to the volume of publicly available data sources. All eQTL and mQTL data obtained, except from the sources eQTLgen, metaBrain and Zeng, et al. (multi ancestry), were already in SMR format and obtained from the Yang Lab’s Data Resource page. ^52,55,56^ The eQTL sources from the Yang Lab include Genotype-Tissue Expression (GTEx) project v8 release, PsychENCODE, and BrainMeta v1 (formerly brain-eMeta).^51,53,54^ The specific tissues measured varied by data source but consisted of NDD-related tissues, which we have defined as brain, nerve, muscle, blood, and liver tissues. Liver was included due to its role in metabolizing medications, toxicity, and potential impacts on clinical trial progress.^69^

mQTLs are genetic variants that affect methylation patterns of CpG sites. mQTL data sources include Brain-mMeta and McRae et al. (2018) which are derived from blood tissues.^54,57^ We included caQTLs - caQTLs alter traits by modifying chromatin structure - data from Bryois et al. (2018) in our analysis. ^59^ Blood tissues have been shown to have high correlation in expression levels with brain tissues, allowing blood tissues to provide a gain of power and ease of use in biomarker studies due to the relative ease of availability of this tissue. ^54^ All genome positions are mapped to the human reference genome build hg19 (GRCh37).

pQTLs are genetic variants associated with protein expression levels. Similarly, to eQTLs and mQTLs, pQTLs can be used as our exposure variable. We obtained pQTL summary statistics data from Yang et al. (2021). ^58^ The pQTLs are from plasma, brain, and cerebrospinal fluid tissues (CSF) from participants with and without AD. Samples are on human reference genome build hg19 (GRCh37). More details on the samples and methods used can be found in the original manuscript.^58^ pQTLs were nominated for inclusion for SMR analysis if they were significant (p < 0.05) in at least one of the three tissues per the original manuscript. We included 453 unique pQTLs across the three tissues: 223 pQTLs from CSF; 159 from plasma; and 77 from the brain.

#### Gene Expression Summarization

We summarized expression ranks for genes of interest within the single cell adult human brain transcriptome data set adult_human_20221007.loom from Siletti et al. (2022).^6^ Using custom R scripts we converted feature counts into TPM (transcripts per million). For a given sample, feature counts were divided by maximum nonredundant intron-removed exon lengths to correct for differences in gene length. Values were then multiplied by a sample-specific constant (10^6^ / T, where T is the sum of length-normalized counts) such that the resulting unitless vector sums to one million. We extracted exon lengths based on annotations from the GTF file used to originally annotate the single cell data (gb_pri_annot.gtf). We calculated the expression percentile rank for genes of interest using the empirical cumulative distribution function and then calculated the mean and median expression percentile rank (EPR) value for each gene across cells of each tested cell type. In order to ease interpretation, we binned the EPR values into 3 classes - off, low, and high (off: EPR < 10, low: 10 < EPR < 90, high: EPR > 90) using the mean snRNA-seq EPR of each gene against cell type.

#### Gene – Gene Networks

Data was obtained from the Open Targets.^39^ Open Targets provides an API to cross reference annotations and relationships on diseases, genes, and drugs. Companion genes were pulled from the SIGnaling Network Open Resource (SIGNOR) database due to the manual curation of gene interactions.^62^

#### Therapeutic Drug Data

Therapeutic drug data was obtained from various sources including Finan, et al. (“druggable genome”), and the Drug Gene Interaction Database. ^48,49^ Druggable genome data were obtained from the supplementary materials in Finan, et al.; The obtained data provided insight on 3000+ potential gene targets with evidence for drug targets or potential targets.^48^ DGIdb drug data (accessed January 2023) were downloaded from the DGIdc online database as files consisting of known gene and drug interactions as well as details such as interaction types and drug categories.

#### Pre-processing

All data pre-processing was carried out using custom scripts for data that was not obtained via the Yang Lab or was missing information such as gene symbols (see Key Resources Table). Preprocessing included gene annotation, BESD format preparation and conversion, and/or calculation of necessary measures such as beta values. Gene annotation was performed as necessary if no gene symbol was provided in the original data source. Annotation was conducted using both the Python biomart package and pyensembl using ENSG IDs. mQTL gene annotation was conducted by obtaining Illumina 450k chip probe data using the R package IlluminaHumanMethylation450kanno.ilmn12.hg19.^50,60,70^

Data sources were converted as necessary into BESD format using the flist method outlined by the Yang Lab. BESD format stores x-QTL summary data in a set of three files - .esi, epi, .besd. More information on the format and how to process data into BESD format can be found on the SMR Yang Lab website listed in the key resources table.

Our multi-ancestry eQTL data originally lacked allele frequency, beta, or standard error values. Missing allele frequencies were obtained using the 1000 Genomes reference panel, from which we derived beta and standard error values using each eQTL’s random effects model z-score, allele frequency and total number of samples from the original study (n = 2,119).

#### Summary data-based Mendelian Randomization

Summary data-based Mendelian Randomization (SMR) is a MR computational tool that uses summary-level data to test if an exposure variable (i.e., gene expression) and outcome (i.e., trait) are causally associated because of a shared causal variant (i.e. instrumental variable).^71^ To discern potentially causal variants from those in linkage disequilibrium with the functional variant, the heterogeneity in dependent instruments (HEIDI) method was implemented using the default version which uses the top 20 SNPs at a locus.^71^ Linkage disequilibrium reference data was obtained from 1000 Genomes phase 3 reference panel.^47^ To increase statistical power, we applied the SMR-multiple exposures (SMR-multi) method, a Bayesian framework for simultaneous testing of multiple traits or exposures on a single outcome while accounting for the correlation between them. SMR and HEIDI analysis were conducted using the SMR software established and maintained by the Yang Lab using all default parameters including those previously detailed.^71,72^

Following SMR, we filtered results to include only protein-coding genes and removed potential associations with no available gene annotations or associations with genes in the major histocompatibility complex (MHC). We used a significance threshold of p_SMR_multi_ < 2.95×10^−6^, corresponding to the Bonferroni-corrected value at α=0.05 for 16,875 protein coding genes tested across all NDD and omic pairs. We then filtered results based on the presence of inferred pleiotropy via the computed HEIDI score (p_HEIDI_ > 0.01 for inclusion in this study).^71^ SNPs were then split on their associated genes status as a therapeutic target or as a non-therapeutic target. After initial processing, analyses were conducted as demonstrated in our workflow diagram (Figure 1 panel a) and explained further in our gene nomination workflow. In total, we tested a total of 186 omic-tissue pairs across six NDD (Supplementary Table S1).

#### Gene Nomination and Drug Target Identification

Gene nomination focused on targets shared by multiple NDD, classifying targets by inferred druggability as described in the introduction. Therapeutic targets were initially nominated using data from DGIdb and Finan, et al..^48,49^ Further target curation was conducted using Open Targets to verify if any approved indications included an NDD thus allowing us to classify drugs into either the *novel* or *known* tiers. Identified network companion genes upstream and downstream of the initial target identified were further categorized into groups based on therapeutic status and approved use in treating any NDD. Our *known* and *difficult* tiers were further investigated using gene co-expression networks via Open Targets interaction annotations through the Signor database. Using Signor identified interactions we identified companion genes, i.e., those manually annotated for their causal relationships with the gene of interest. We additionally searched known therapeutics that target any identified companion genes to potentially identify proxy gene targets thus expanding the net for drug discovery and repurposing. We implemented custom python scripts in order to query Open Targets’ API to extract relevant annotations for this workflow.

## Notes

### Funding Statement

We would like to thank all of the subjects who donated their time and biological samples to the studies that are reflected in these analyses. This research was supported in part by the Intramural Research Program of the National Institutes of Health (National Institute on Aging and National Institute of Neurological Disorders and Stroke; project numbers: 1ZIAAG00935, 1ZIANS003154, Z01-AG000949-02). This study used the high-performance computational capabilities of the Biowulf Linux cluster at the National Institutes of Health (http://hpc.nih.gov).

### Author Declarations

The study used ONLY openly available human data that were originally located at https://nih-card-ndd-smr-home-syboky.streamlit.app/About

### Summary of Updates

Author added

## References

1. Van Dyck, C.H., Swanson, C.J., Aisen, P., Bateman, R.J., Chen, C., Gee, M., Kanekiyo, M., Li, D., Reyderman, L., Cohen, S., et al. (2023). Lecanemab in Early Alzheimer’s Disease. New England Journal of Medicine 388, 9–21. 10.1056/nejmoa2212948.

2. Morant, A.V., Jagalski, V., and Vestergaard, H.T. (2019). Labeling of Disease-Modifying Therapies for Neurodegenerative Disorders. Frontiers in Medicine 6.

3. Organization, W.H. Dementia. https://www.who.int/news-room/fact-sheets/detail/dementia.

4. King, E.A., Davis, J.W., and Degner, J.F. (2019). Are drug targets with genetic support twice as likely to be approved? Revised estimates of the impact of genetic support for drug mechanisms on the probability of drug approval. PLOS Genetics 15, e1008489. 10.1371/journal.pgen.1008489.

5. Paulsen, J.S., Nance, M., Kim, J.-I., Carlozzi, N.E., Panegyres, P.K., Erwin, C., Goh, A., McCusker, E., and Williams, J.K. (2013). A review of quality of life after predictive testing for and earlier identification of neurodegenerative diseases. Progress in Neurobiology 110, 2–28. 10.1016/j.pneurobio.2013.08.003.

6. Siletti, K., Hodge, R., Mossi Albiach, A., Hu, L., Lee, K.W., Lönnerberg, P., Bakken, T., Ding, S.-L., Clark, M., Casper, T., et al. (2022). Transcriptomic diversity of cell types across the adult human brain. Cold Spring Harbor Laboratory.

7. Alvarado, C.X., Weller, C.A., Johnson, N.L., Leonard, H.L., Singleton, A.B., Reed, X., Blauewendraat, C., and Nalls, M.A. (2023). Human brain single nucleus cell type enrichments in neurodegenerative diseases. Cold Spring Harbor Laboratory.

8. Seo, J., and Park, M. (2020). Molecular crosstalk between cancer and neurodegenerative diseases. Cellular and Molecular Life Sciences 77, 2659–2680. 10.1007/s00018-019-03428-3.

9. Wilson, D.M., Cookson, M.R., Van Den Bosch, L., Zetterberg, H., Holtzman, D.M., and Dewachter, I. (2023). Hallmarks of neurodegenerative diseases. Cell 186, 693–714. https://doi.org/10.1016/j.cell.2022.12.032.

10. Zhou, F., and Wang, D. (2017). The associations between the MAPT polymorphisms and Alzheimer’s disease risk: a meta-analysis. Oncotarget 8, 43506–43520. 10.18632/oncotarget.16490.

11. Tobin, J.E., Latourelle, J.C., Lew, M.F., Klein, C., Suchowersky, O., Shill, H.A., Golbe, L.I., Mark, M.H., Growdon, J.H., Wooten, G.F., et al. (2008). Haplotypes and gene expression implicate the MAPT region for Parkinson disease: The GenePD Study. Neurology 71, 28–34. 10.1212/01.wnl.0000304051.01650.23.

12. Majounie, E., Cross, W., Newsway, V., Dillman, A., Vandrovcova, J., Morris, C.M., Nalls, M.A., Ferrucci, L., Owen, M.J., O’Donovan, M.C., et al. (2013). Variation in tau isoform expression in different brain regions and disease states. Neurobiology of Aging 34, 1922.e1927–1922.e1921. 10.1016/j.neurobiolaging.2013.01.017.

13. Lin, M.K., and Farrer, M.J. (2014). Genetics and genomics of Parkinson’s disease. Genome Medicine 6, 48. 10.1186/gm566.

14. Jun, G., Ibrahim-Verbaas, C.A., Vronskaya, M., Lambert, J.C., Chung, J., Naj, A.C., Kunkle, B.W., Wang, L.S., Bis, J.C., Bellenguez, C., et al. (2016). A novel Alzheimer disease locus located near the gene encoding tau protein. Molecular Psychiatry 21, 108–117. 10.1038/mp.2015.23.

15. Patel, D., Zhang, X., Farrell, J.J., Chung, J., Stein, T.D., Lunetta, K.L., and Farrer, L.A. (2021). Cell-type-specific expression quantitative trait loci associated with Alzheimer disease in blood and brain tissue. Translational Psychiatry 11. 10.1038/s41398-021-01373-z.

16. Futch, H.S., Croft, C.L., Truong, V.Q., Krause, E.G., and Golde, T.E. (2017). Targeting psychologic stress signaling pathways in Alzheimer’s disease. Molecular Neurodegeneration 12. 10.1186/s13024-017-0190-z.

17. Zhao, B., Shan, Y., Yang, Y., Yu, Z., Li, T., Wang, X., Luo, T., Zhu, Z., Sullivan, P., Zhao, H., et al. (2021). Transcriptome-wide association analysis of brain structures yields insights into pleiotropy with complex neuropsychiatric traits. Nature Communications 12. 10.1038/s41467-021-23130-y.

18. Madrid, L., Labrador, S.C., González-Pérez, A., and Sáez, M.E. (2021). Integrated Genomic, Transcriptomic and Proteomic Analysis for Identifying Markers of Alzheimer’s Disease. Diagnostics 11, 2303. 10.3390/diagnostics11122303.

19. Sheikh, B.N., Guhathakurta, S., and Akhtar, A. (2019). The nonDspecific lethal NSL complex at the crossroads of transcriptional control and cellular homeostasis. EMBO reports 20, e47630. 10.15252/embr.201847630.

20. Linda, K., Lewerissa, E.I., Verboven, A.H.A., Gabriele, M., Frega, M., Klein Gunnewiek, T.M., Devilee, L., Ulferts, E., Hommersom, M., Oudakker, A., et al. (2022). Imbalanced autophagy causes synaptic deficits in a human model for neurodevelopmental disorders. Autophagy 18, 423–442. 10.1080/15548627.2021.1936777.

21. Button, R.W., Roberts, S.L., Willis, T.L., Hanemann, C.O., and Luo, S. (2017). Accumulation of autophagosomes confers cytotoxicity. Journal of Biological Chemistry 292, 13599–13614. 10.1074/jbc.m117.782276.

22. Son, S.M., Park, S.J., Fernandez-Estevez, M., and Rubinsztein, D.C. (2021). Autophagy regulation by acetylation—implications for neurodegenerative diseases. Experimental & Molecular Medicine 53, 30–41. 10.1038/s12276-021-00556-4.

23. Lingling, D., Miaomiao, Q., Yili, L., Hongyun, H., and Yihao, D. (2022). Attenuation of histone H4 lysine 16 acetylation (H4K16ac) elicits a neuroprotection against ischemic stroke by alleviating the autophagic/lysosomal dysfunction in neurons at the penumbra. Brain Research Bulletin 184, 24–33. https://doi.org/10.1016/j.brainresbull.2022.03.013.

24. Füllgrabe, J., Lynch-Day, M.A., Heldring, N., Li, W., Struijk, R.B., Ma, Q., Hermanson, O., Rosenfeld, M.G., Klionsky, D.J., and Joseph, B. (2013). The histone H4 lysine 16 acetyltransferase hMOF regulates the outcome of autophagy. Nature 500, 468–471. 10.1038/nature12313.

25. Wiesel-Motiuk, N., and Assaraf, Y.G. (2020). The key roles of the lysine acetyltransferases KAT6A and KAT6B in physiology and pathology. Drug Resistance Updates 53, 100729. https://doi.org/10.1016/j.drup.2020.100729.

26. Wang, C., Telpoukhovskaia, M.A., Bahr, B.A., Chen, X., and Gan, L. (2018). Endo-lysosomal dysfunction: a converging mechanism in neurodegenerative diseases. Current Opinion in Neurobiology 48, 52–58. https://doi.org/10.1016/j.conb.2017.09.005.

27. Kragh, C.L., Ubhi, K., Wyss-Corey, T., and Masliah, E. (2012). Autophagy in Dementias. Brain Pathology 22, 99–109. 10.1111/j.1750-3639.2011.00545.x.

28. David, Popovic, D., Gubas, A., Terawaki, S., Suzuki, H., Stadel, D., Fraser, Diana, Bhogaraju, S., Maddi, K., et al. (2015). PLEKHM1 Regulates Autophagosome-Lysosome Fusion through HOPS Complex and LC3/GABARAP Proteins. Molecular Cell 57, 39–54. 10.1016/j.molcel.2014.11.006.

29. Lichtenthaler, S.F., Lemberg, M.K., and Fluhrer, R. (2018). Proteolytic ectodomain shedding of membrane proteins in mammals—hardware, concepts, and recent developments. The EMBO Journal 37. 10.15252/embj.201899456.

30. Chen, F., Chen, H., Jia, Y., Lu, H., Tan, Q., and Zhou, X. (2020). miRD149D5p inhibition reduces Alzheimer’s disease β-amyloid generation in 293/APPsw cells by upregulating H4K16ac via KAT8. Experimental and Therapeutic Medicine 20, 1–1. 10.3892/etm.2020.9216.

31. Erekat, N.S. (2018). Apoptosis and its Role in Parkinson’s Disease. In (Codon Publications), pp. 65–82. 10.15586/codonpublications.parkinsonsdisease.2018.ch4.

32. Goel, P., Chakrabarti, S., Goel, K., Bhutani, K., Chopra, T., and Bali, S. (2022). Neuronal cell death mechanisms in Alzheimer’s disease: An insight. Front Mol Neurosci 15, 937133. 10.3389/fnmol.2022.937133.

33. Van Den Bosch, T., Leus, N.G.J., Wapenaar, H., Boichenko, A., Hermans, J., Bischoff, R., Haisma, H.J., and Dekker, F.J. (2017). A 6-alkylsalicylate histone acetyltransferase inhibitor inhibits histone acetylation and pro-inflammatory gene expression in murine precision-cut lung slices. Pulmonary Pharmacology & Therapeutics 44, 88–95. 10.1016/j.pupt.2017.03.006.

34. Dekker, F.J., van den Bosch, T., and Martin, N.I. (2014). Small molecule inhibitors of histone acetyltransferases and deacetylases are potential drugs for inflammatory diseases. Drug Discovery Today 19, 654–660. https://doi.org/10.1016/j.drudis.2013.11.012.

35. Baird, D.A., Liu, J.Z., Zheng, J., Sieberts, S.K., Perumal, T., Elsworth, B., Richardson, T.G., Chen, C.-Y., Carrasquillo, M.M., Allen, M., et al. (2021). Identifying drug targets for neurological and psychiatric disease via genetics and the brain transcriptome. PLOS Genetics 17, e1009224. 10.1371/journal.pgen.1009224.

36. Li, Y.I., Wong, G., Humphrey, J., and Raj, T. (2019). Prioritizing Parkinson’s disease genes using population-scale transcriptomic data. Nature Communications 10. 10.1038/s41467-019-08912-9.

37. Diaz-Ortiz, M.E., Seo, Y., Posavi, M., Carceles Cordon, M., Clark, E., Jain, N., Charan, R., Gallagher, M.D., Unger, T.L., Amari, N., et al. (2022). GPNMB confers risk for Parkinson’s disease through interaction with α-synuclein. Science 377, eabk0637. 10.1126/science.abk0637.

38. Nalls, M.A., Pankratz, N., Lill, C.M., Do, C.B., Hernandez, D.G., Saad, M., Destefano, A.L., Kara, E., Bras, J., Sharma, M., et al. (2014). Large-scale meta-analysis of genome-wide association data identifies six new risk loci for Parkinson’s disease. Nature Genetics 46, 989–993. 10.1038/ng.3043.

39. Ochoa, D., Hercules, A., Carmona, M., Suveges, D., Baker, J., Malangone, C., Lopez, I., Miranda, A., Cruz-Castillo, C., Fumis, L., et al. (2023). The next-generation Open Targets Platform: reimagined, redesigned, rebuilt. Nucleic Acids Research 51, D1353–D1359. 10.1093/nar/gkac1046.

40. Pajares, M., A, I.R., Manda, G., Boscá, L., and Cuadrado, A. (2020). Inflammation in Parkinson’s Disease: Mechanisms and Therapeutic Implications. Cells 9. 10.3390/cells9071687.

41. Marogianni, C., Sokratous, M., Dardiotis, E., Hadjigeorgiou, G.M., Bogdanos, D., and Xiromerisiou, G. (2020). Neurodegeneration and Inflammation—An Interesting Interplay in Parkinson’s Disease. International Journal of Molecular Sciences 21, 8421. 10.3390/ijms21228421.

42. Matthews, D.C., Mao, X., Dowd, K., Tsakanikas, D., Jiang, C.S., Meuser, C., Andrews, R.D., Lukic, A.S., Lee, J., Hampilos, N., et al. (2021). Riluzole, a glutamate modulator, slows cerebral glucose metabolism decline in patients with Alzheimer’s disease. Brain 144, 3742–3755. 10.1093/brain/awab222.

43. Wu, H.-C., Chen, C.-M., Chen, Y.-C., Fung, H.-C., Chang, K.-H., and Wu, Y.-R. (2018). DLG2, but not TMEM229B, GPNMB, and ITGA8 polymorphism, is associated with Parkinson’s disease in a Taiwanese population. Neurobiology of Aging 64, 158.e151–158.e156. https://doi.org/10.1016/j.neurobiolaging.2017.11.016.

44. Xu, Y., Chen, Y., Ou, R., Wei, Q.-Q., Cao, B., Chen, K., and Shang, H.-F. (2016). No association of GPNMB rs156429 polymorphism with Parkinson’s disease, amyotrophic lateral sclerosis and multiple system atrophy in Chinese population. Neuroscience Letters 622, 113–117. https://doi.org/10.1016/j.neulet.2016.04.060.

45. Mie, R., Sara, B.-C., Mary, B.M., Oluwadamilola, O., Peter Wild, C., Oladunni, A., Kristin, S.L., Sani, A., Charles, A., Dan, V., et al. (2023). Genome-wide Association Identifies Novel Etiological Insights Associated with Parkinson’s Disease in African and African Admixed Populations. medRxiv, 2023.2005.2005.23289529. 10.1101/2023.05.05.23289529.

46. Program, N.I.o.H.A.o.U.R. All of Us Research Hub. https://www.researchallofus.org/.

47. Auton, A., Abecasis, G.R., Altshuler, D.M., Durbin, R.M., Abecasis, G.R., Bentley, D.R., Chakravarti, A., Clark, A.G., Donnelly, P., Eichler, E.E., et al. (2015). A global reference for human genetic variation. Nature 526, 68–74. 10.1038/nature15393.

48. Finan, C., Gaulton, A., Kruger, F.A., Lumbers, R.T., Shah, T., Engmann, J., Galver, L., Kelley, R., Karlsson, A., Santos, R., et al. (2017). The druggable genome and support for target identification and validation in drug development. Science Translational Medicine 9, eaag1166. 10.1126/scitranslmed.aag1166.

49. Freshour, S.L., Kiwala, S., Cotto, K.C., Coffman, A.C., McMichael, J.F., Song, J.J., Griffith, M., Griffith, Obi L., and Wagner, A.H. (2021). Integration of the Drug–Gene Interaction Database (DGIdb 4.0) with open crowdsource efforts. Nucleic Acids Research 49, D1144–D1151. 10.1093/nar/gkaa1084.

50. Hansen, K.D. (2016). IlluminaHumanMethylation450kanno.ilmn12.hg19: Annotation for Illumina’s 450k methylation arrays. https://bioconductor.org/packages/release/data/annotation/html/IlluminaHumanMethylation450kanno.ilmn12.hg19.html.

51. The, G.C., Aguet, F., Anand, S., Ardlie, K.G., Gabriel, S., Getz, G.A., Graubert, A., Hadley, K., Handsaker, R.E., Huang, K.H., et al. (2020). The GTEx Consortium atlas of genetic regulatory effects across human tissues. Science 369, 1318–1330. 10.1126/science.aaz1776.

52. Võsa, U., Claringbould, A., Westra, H.-J., Bonder, M.J., Deelen, P., Zeng, B., Kirsten, H., Saha, A., Kreuzhuber, R., Yazar, S., et al. (2021). Large-scale cis- and trans-eQTL analyses identify thousands of genetic loci and polygenic scores that regulate blood gene expression. Nature Genetics 53, 1300–1310. 10.1038/s41588-021-00913-z.

53. Wang, D., Liu, S., Warrell, J., Won, H., Shi, X., Navarro, F.C.P., Clarke, D., Gu, M., Emani, P., Yang, Y.T., et al. (2018). Comprehensive functional genomic resource and integrative model for the human brain. Science 362, eaat8464. 10.1126/science.aat8464.

54. Qi, T., Wu, Y., Zeng, J., Zhang, F., Xue, A., Jiang, L., Zhu, Z., Kemper, K., Yengo, L., Zheng, Z., et al. (2018). Identifying gene targets for brain-related traits using transcriptomic and methylomic data from blood. Nature Communications 9. 10.1038/s41467-018-04558-1.

55. De Klein, N., Tsai, E.A., Vochteloo, M., Baird, D., Huang, Y., Chen, C.-Y., Van Dam, S., Oelen, R., Deelen, P., Bakker, O.B., et al. (2023). Brain expression quantitative trait locus and network analyses reveal downstream effects and putative drivers for brain-related diseases. Nature Genetics 55, 377–388. 10.1038/s41588-023-01300-6.

56. Zeng, B., Bendl, J., Kosoy, R., Fullard, J.F., Hoffman, G.E., and Roussos, P. (2022). Multi-ancestry eQTL meta-analysis of human brain identifies candidate causal variants for brain-related traits. Nature Genetics 54, 161–169. 10.1038/s41588-021-00987-9.

57. McRae, A.F., Marioni, R.E., Shah, S., Yang, J., Powell, J.E., Harris, S.E., Gibson, J., Henders, A.K., Bowdler, L., Painter, J.N., et al. (2018). Identification of 55,000 Replicated DNA Methylation QTL. Scientific Reports 8. 10.1038/s41598-018-35871-w.

58. Yang, C., Farias, F.H.G., Ibanez, L., Suhy, A., Sadler, B., Fernandez, M.V., Wang, F., Bradley, J.L., Eiffert, B., Bahena, J.A., et al. (2021). Genomic atlas of the proteome from brain, CSF and plasma prioritizes proteins implicated in neurological disorders. Nature Neuroscience 24, 1302–1312. 10.1038/s41593-021-00886-6.

59. Bryois, J., Garrett, M.E., Song, L., Safi, A., Giusti-Rodriguez, P., Johnson, G.D., Shieh, A.W., Buil, A., Fullard, J.F., Roussos, P., et al. (2018). Evaluation of chromatin accessibility in prefrontal cortex of individuals with schizophrenia. Nature Communications 9. 10.1038/s41467-018-05379-y.

60. Openvax (2017). PyEnsembl. https://github.com/openvax/pyensembl.

61. Kent, W.J., Sugnet, C.W., Furey, T.S., Roskin, K.M., Pringle, T.H., Zahler, A.M., Haussler, and David (2002). The Human Genome Browser at UCSC. Genome Research 12, 996–1006. 10.1101/gr.229102.

62. Lo Surdo, P., Iannuccelli, M., Contino, S., Castagnoli, L., Licata, L., Cesareni, G., and Perfetto, L. (2023). SIGNOR 3.0, the SIGnaling network open resource 3.0: 2022 update. Nucleic Acids Research 51, D631–D637. 10.1093/nar/gkac883.

63. Bellenguez, C., Küçükali, F., Jansen, I.E., Kleineidam, L., Moreno-Grau, S., Amin, N., Naj, A.C., Campos-Martin, R., Grenier-Boley, B., Andrade, V., et al. (2022). New insights into the genetic etiology of Alzheimer’s disease and related dementias. Nature Genetics 54, 412–436. 10.1038/s41588-022-01024-z.

64. Nicolas, A., Kenna, K.P., Renton, A.E., Ticozzi, N., Faghri, F., Chia, R., Dominov, J.A., Kenna, B.J., Nalls, M.A., Keagle, P., et al. (2018). Genome-wide Analyses Identify KIF5A as a Novel ALS Gene. Neuron 97, 1268–1283.e1266. 10.1016/j.neuron.2018.02.027.

65. Pottier, C., Ren, Y., Perkerson, R.B., Baker, M., Jenkins, G.D., Van Blitterswijk, M., Dejesus-Hernandez, M., Van Rooij, J.G.J., Murray, M.E., Christopher, E., et al. (2019). Genome-wide analyses as part of the international FTLD-TDP whole-genome sequencing consortium reveals novel disease risk factors and increases support for immune dysfunction in FTLD. Acta Neuropathologica 137, 879–899. 10.1007/s00401-019-01962-9.

66. Chia, R., Sabir, M.S., Bandres-Ciga, S., Saez-Atienzar, S., Reynolds, R.H., Gustavsson, E., Walton, R.L., Ahmed, S., Viollet, C., Ding, J., et al. (2021). Genome sequencing analysis identifies new loci associated with Lewy body dementia and provides insights into its genetic architecture. Nature Genetics 53, 294–303. 10.1038/s41588-021-00785-3.

67. Nalls, M.A., Blauwendraat, C., Vallerga, C.L., Heilbron, K., Bandres-Ciga, S., Chang, D., Tan, M., Kia, D.A., Noyce, A.J., Xue, A., et al. (2019). Identification of novel risk loci, causal insights, and heritable risk for Parkinson’s disease: a meta-analysis of genome-wide association studies. The Lancet Neurology 18, 1091–1102. 10.1016/s1474-4422(19)30320-5.

68. Höglinger, G.U., Melhem, N.M., Dickson, D.W., Sleiman, P.M.A., Wang, L.-S., Klei, L., Rademakers, R., De Silva, R., Litvan, I., Riley, D.E., et al. (2011). Identification of common variants influencing risk of the tauopathy progressive supranuclear palsy. Nature Genetics 43, 699–705. 10.1038/ng.859.

69. Vaja, R., and Rana, M. (2020). Drugs and the liver. Anaesthesia & Intensive Care Medicine 21, 517–523. 10.1016/j.mpaic.2020.07.001.

70. Briois, S. Biomart. https://github.com/sebriois/biomart.

71. Zhu, Z., Zhang, F., Hu, H., Bakshi, A., Robinson, M.R., Powell, J.E., Montgomery, G.W., Goddard, M.E., Wray, N.R., Visscher, P.M., and Yang, J. (2016). Integration of summary data from GWAS and eQTL studies predicts complex trait gene targets. Nature Genetics 48, 481–487. 10.1038/ng.3538.

72. Wu, Y., Zeng, J., Zhang, F., Zhu, Z., Qi, T., Zheng, Z., Lloyd-Jones, L.R., Marioni, R.E., Martin, N.G., Montgomery, G.W., et al. (2018). Integrative analysis of omics summary data reveals putative mechanisms underlying complex traits. Nature Communications 9. 10.1038/s41467-018-03371-0.

