## Supplemental File for "omicSynth: an Open Multi-omic Community Resource for Identifying Druggable Targets across Neurodegenerative Diseases"

### Supplementary Tables Legend

**Table S1: Summary of SMR data mining across diseases.** This table provides a summary of the number of unique genes, genes found to be significant in liver eQTL tissue, eQTL specific genes, number of genes replicated in multi ancestry eQTL data, total number of gene found to be therapeutic/druggable, the percentage of therapeutic genes, total number of non-therapeutic/non-druggable genes, and the percentage of non-therapeutic genes for each diseases. This data is provided at 4 levels, an overall summary of all tested genes and three significance thresholds: $p_{SMR\_multi} < 0.05$ & $p_{HEIDI} > 0.01$, $p_{SMR\_multi} < 2.95 \times 10^{-6}$ (testing all protein coding genes) & $p_{HEIDI} > 0.01$, and $p_{SMR\_multi} < 1.58 \times 10^{-8}$ (testing all protein coding genes across all omics) & $p_{HEIDI} > 0.01$.

**Table S2:All unfiltered SMR association summary statistics.** Table provides a link to data files for download.

**Table S3: Extended data for candidate genes for multiple neurodegenerative diseases (Table 1).** This table provides extended data for genes with functional inferences passing multiple test correction at a multi-SNP SMR $P < 2.95 \times 10^{-6}$ for multiple neurodegenerative diseases. We provide details for the most significant *-omic association detected as well as the most significant single SNP in that set of gene level associations. Additionally, we provide all the omics and diseases in which a given gene has significant associations.

**Table S4: Summary statistics for all significant gene associations.** This table provides summary statistics and SMR results for candidate genes with associations passing multiple test correction at a multi-SNP SMR P < 2.95E-06.

**Table S5: Extended Summaries of candidate genes in three neurodegenerative diseases.** This table provides extended data on candidate genes passing multiple test correction at a multi-SNP SMRP < 2.95E-06 for three neurodegenerative diseases. We provide summary statistics for each significant association.

**Table S6: Extended Summaries of candidate genes in two neurodegenerative diseases.** This table provides extended data on candidate genes passing multiple test correction at a multi-SNP SMRP < 2.95E-06 for two neurodegenerative diseases. We provide summary statistics for each significant association.

**Table S7: Summary statistics for candidate genes replicated in multi ancestry data.** This table provides summary statistics for all replicated candidate genes’ significant associations across all *-omics.

**Table S8: Summary statistics for all novel class genes.** This table provides summary statistics of candidate genes classified as *novel,* meaning the gene is considered druggable but does not have NDD specific therapeutics that target the gene. Significant associations for each gene are provided.

**Table S9: Summary statistics for all known class genes.** This table provides summary statistics of candidate genes classified as *known,* meaning that the gene has therapeutics that are approved for use in at least one NDD. We provide all significant associations for each *known* gene.

**Table S10: Summaries of the novel target class of genes including network members.** This table lists all candidate genes that fall into the novel tier in our druggable gene classification scheme. For each gene we provide a summary of how many omics a gene has significant associations in and in which diseases. We identify companion genes for each gene using the Signor database and in bold are genes that have been found to have significant associations in our SMR results (multi-SNP SMRP < 2.95E-06).

**Table S11: Summary statistics for all difficult class genes.** This table provides summary statistics of candidate genes classified as *difficult,* meaning the gene is not currently considered druggable and no therapeutics target the gene.

**Table S12: Summaries of the difficult target class of genes including network members.**This table lists all candidate genes that fall into the difficult tier in our druggable gene classification scheme. For each gene we provide a summary of how many omics a gene has significant associations in and in which diseases. We identify companion genes for each gene using the Signor database and in bold are genes that have been found to have significant associations in our SMR results (multi-SNP SMRP < 2.95E-06). We provide additional information regarding potential liver toxicity issues by cross referencing companion genes against significant associations in the liver omic.

**Table S13: Mean expression percentile ranks in single cell data for multi-NDD gene targets.** This table provides calculated mean expression rank values using snRNA-seq data from Silleti et al. (2022). Mean values for each gene-cell type were obtained by using scRNA-seq values for all cell type specific individual cells for a specific gene.

**Table S14: Median expression percentile ranks in single cell data for multi-NDD gene targets.** This table provides calculated median expression rank values using snRNA-seq data from Silleti et al. (2022). Median values for each gene-cell type were obtained by using scRNA-seq values for all cell type specific individual cells for a specific gene.

**Table S15: Comparison of GPNMB across multiple genetic ancestries.** This table provides basic summary statistics of our GPNMB significant associations in comparison to other GPNMB summary statistics. We summarize data in African American/African admixed, Chinese, and Taiwanese genetic ancestries to confirm that our multi ancestry association direction was consistent with previous findings in other non-European ancestries.

### Supplementary Figure(s)

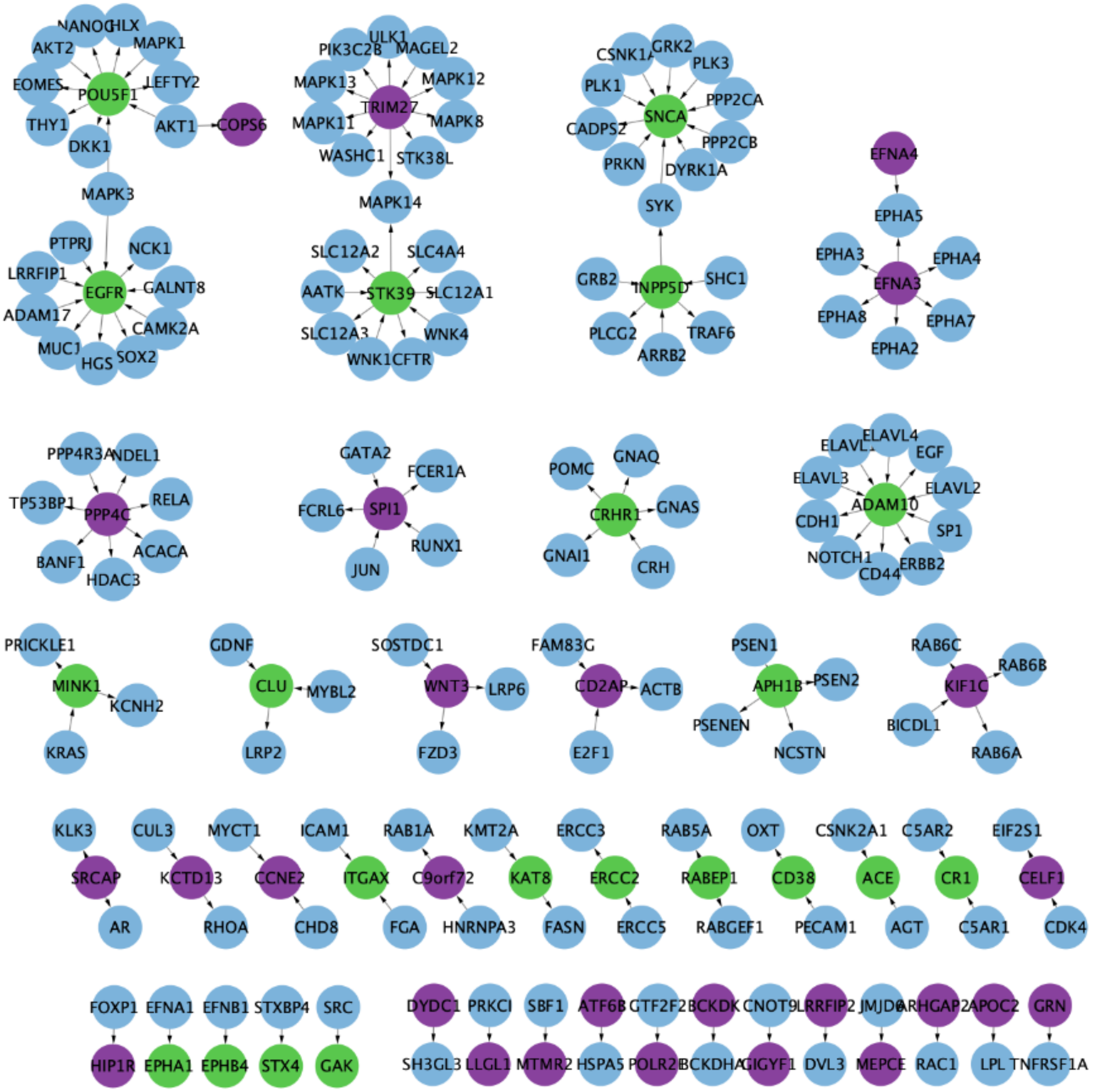


**Supplementary Figure 1**: Network Visualization of *Novel* and *Difficult* genes companion genes Graph network visualization of both *novel* (green nodes) and *difficult* (purple nodes) genes and their SIGNOR curated partners (blue nodes). The direction of connecting arrows indicates interaction from regulator to target.

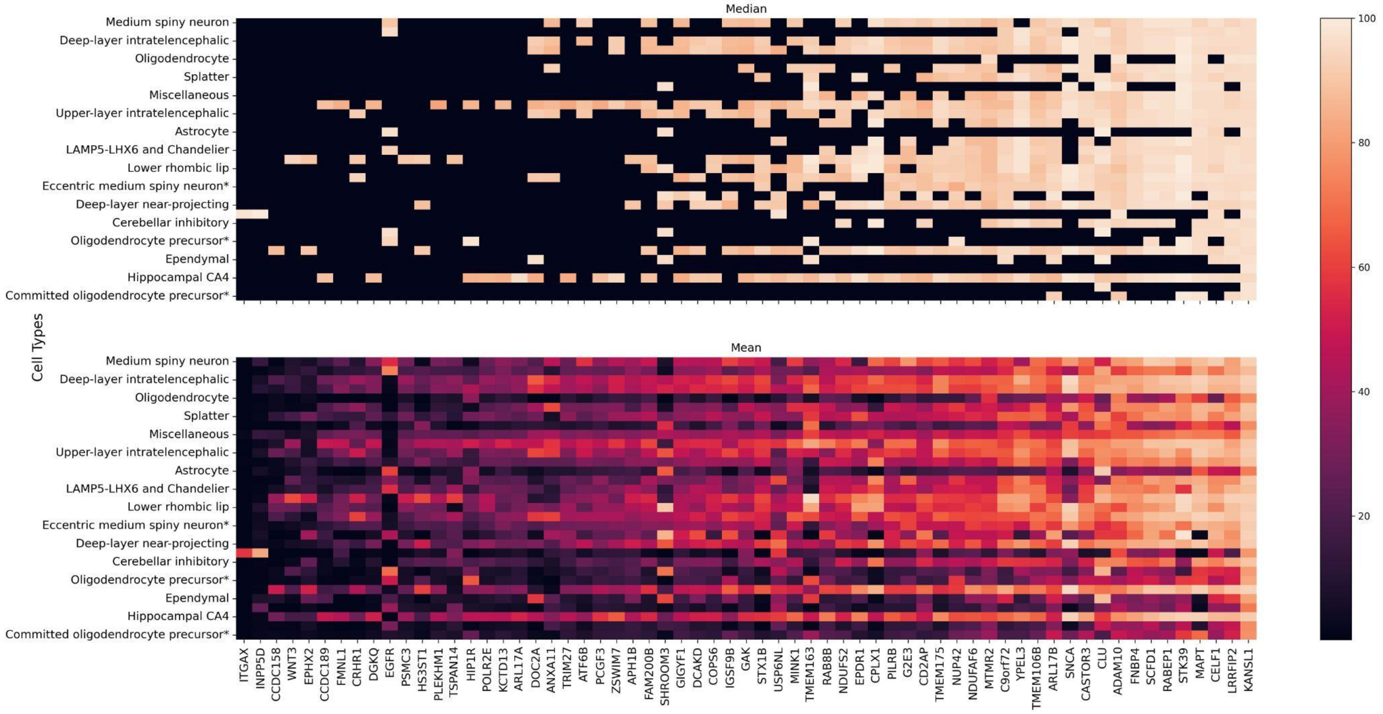


**Supplementary Figure 1: scRNA-seq expression for significant genes ($p_{SMR\_multi}$ < 2.95×10-6 and $p_{HEIDI}$ > 0.01).** Heatmaps were used to illustrate both the median (top plot) and mean (bottom plot) expression percentile rank for each gene-celltype combination using significant genes and disease relevant cell types.

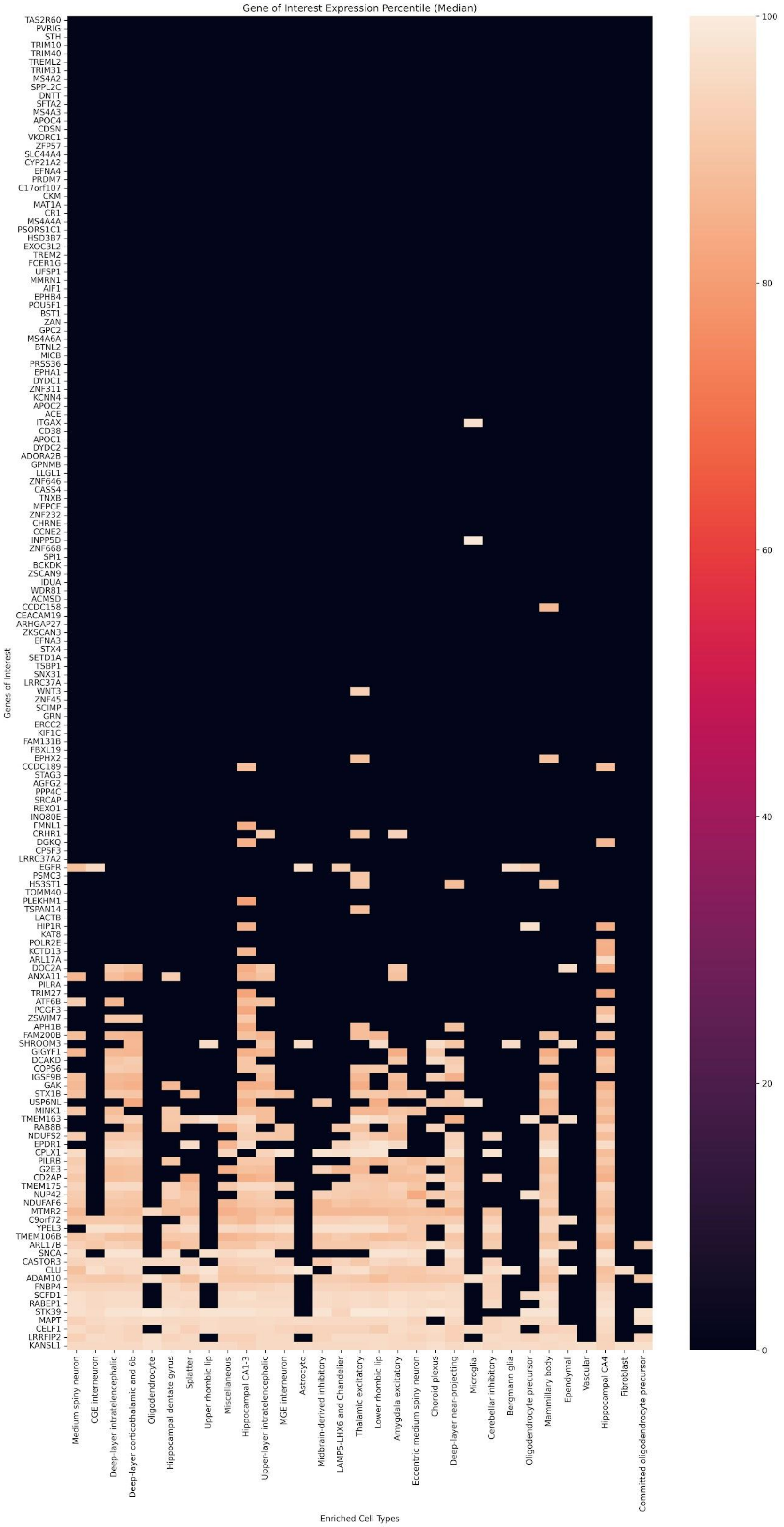


**Supplementary Figure 2: Median scRNA-seq expression for all significant genes.** Heatmap illustration of the median expression percentile rank value for each gene-celltype combination.

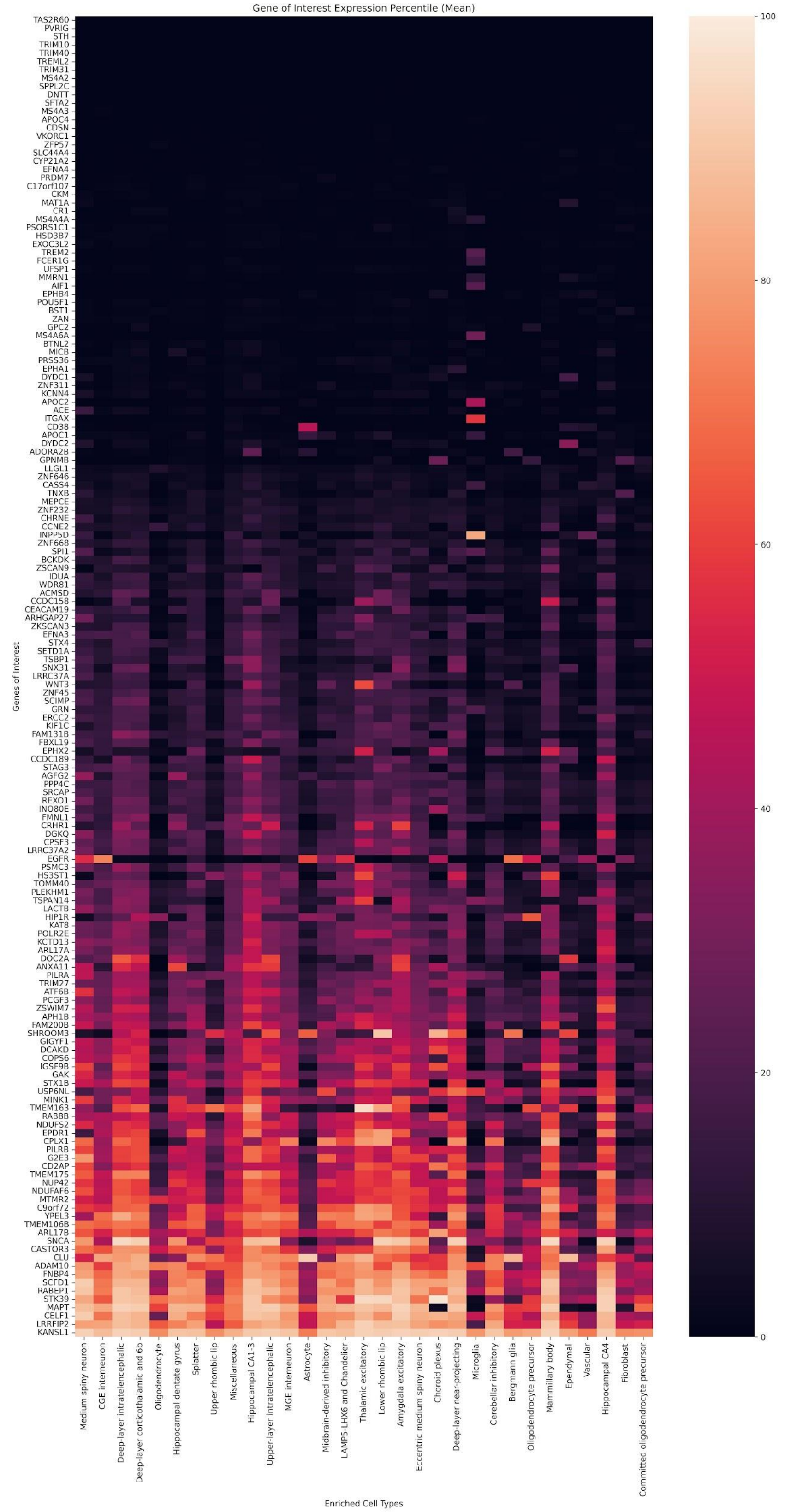


**Supplementary Figure 3: Mean scRNA-seq expression for all significant genes.** Heatmap illustration of the mean expression percentile rank value for each gene-celltype combination.
